# Voxel-level forecast system for lesion development in patients with COVID-19

**DOI:** 10.1101/2020.12.17.20248377

**Authors:** Cheng Jin, Yongjie Duan, Yukun Cao, Jinyang Yu, Zhanwei Xu, Weixiang Chen, Xiaoyu Han, Jia Liu, Jie Zhou, Heshui Shi, Jianjiang Feng

## Abstract

The global spread of COVID-19 seriously endangers human health and even lives. By predicting patients’ individualized disease development and further performing intervention in time, we may rationalize scarce medical resources and reduce mortality. Based on 1337 multi-stage (≥3) high-resolution chest computed tomography (CT) images of 417 infected patients from three centers in the epidemic area, we proposed a random forest + cellular automata (RF+CA) model to forecast voxel-level lesion development of patients with COVID-19. The model showed a promising prediction performance (Dice similarity coefficient [DSC] = 71.1%, Kappa coefficient = 0.612, Figure of Merit [FoM] = 0.257, positional accuracy [PA] = 3.63) on the multicenter dataset. Using this model, multiple driving factors for the development of lesions were determined, such as distance to various interstitials in the lung, distance to the pleura, etc. The driving processes of these driving factors were further dissected and explained in depth from the perspective of pathophysiology, to explore the mechanism of individualized development of COVID-19 disease. The complete codes of the forecast system are available at *https://github.com/keyunj/VVForecast_covid19*.

## Introduction

The outbreak of COVID-19 has put enormous pressure on global health services. The World Health Organization has classified COVID-19 as a pandemic^1^. The development of infected patient’s condition is rapid and unpredictable. Some patients can progress to a severe or critical state within 48 hours^2^. About 15% of COVID-19 infections in Wuhan, China, were reported to be severe^3^. Recent reports indicated that case fatality rate is of over 55% among critical cases, which rises sharply with age and underlying comorbid diseases^4^. The progression to severe disease has brought great pressure to medical services, resulting in the failure of timely allocation of intensive care resources, which further brings great risks to patients’ lives. Early and accurate assessment of disease dynamics and prediction of disease trends is critical, since timely intervention treatment can effectively reduce the occurrence of severe diseases and improve the prognosis of patients.

Various laboratory indicators have been used to assess the severity of the patients with infectious pneumonia and to guide clinical interventions, such as neutrophils lymphocyte ratio (NLR)^5-7^, lactic acid and D-dimer level^8,9^ etc. Researchers also adopt acute physiology and chronic health evaluation (APACHE-II) score system to predict the prognosis of acute respiratory distress syndrome (ARDS)^9^. These existing methods attempt to explore the relevance between some biomarkers and the severity of the disease on a population level, which are insufficient to predict the specific development of pulmonary lesions.

Chest imaging, especially CT scan, is important for the diagnosis and management of COVID-19 patients. High resolution CT scan (HRCT) objectively demonstrates pulmonary lesions and enables us to better understand the pathogenesis of the disease. Various studies have been conducted to diagnose, detect and segment the COVID-19 lesions using CT scans^10-14^. Also, through serial CT examinations, the evolution of the disease can be observed and understood^15-18^. It is of more clinical significance to predict future trend of lesion development than to analyze the development in the past^16,19,20^. Up to now, however, chest CT based prognostic models mainly focus on predicting whether a patient will progress to severe disease or not^7,16^, which is a two-class classification problem. The voxel-level development prediction of pneumonia lesions has not been explored in previous studies. It is a much more challenging task due to limited number of CT examinations, large and unequal examination intervals, rapid and changeable lesion development, etc. Besides, the development of pulmonary infection varies significantly from patient to patient, and even from lobe from lobe.

In this study, we retrospectively analyzed evolution rules of lesions based on multi-stage chest CT images of 417 COVID-19 patients from three centers and proposed an artificial intelligence (AI) system to forecast the development of lesions in the three-dimensional space. The problem caused by uneven intervals between multi-stage CT examinations was overcome by the fusion of spatio-temporal information. Additionally, by measuring the driving factors of lesion evolution, the pathogenesis of COVID-19 was explained, and the time course of lesion absorption and expansion was tracked and analyzed.

## Result

### Datasets for system development and evaluation

A total of 2877 3D CT volumes of 1505 patients with COVID-19 were collected from three centers in Wuhan from February 5 to April 27, 2020. Because the forecast system was trained by the dynamic changes between the 1^st^ and 2^nd^ stage CT volumes and then validated using the 3^rd^ or later stage, only patients with at least three stages of CT volumes were selected. We screened out 636 patients with single stage and 452 patients with 2 stages, and finally obtained 417 patients with at least 3 stages. Therefore, 1337 CT volumes from 417 patients were used for evaluating our proposed forecast system for lesion development (detailly described in Methods and **Extended data | Table 1**).

**Table 1.**
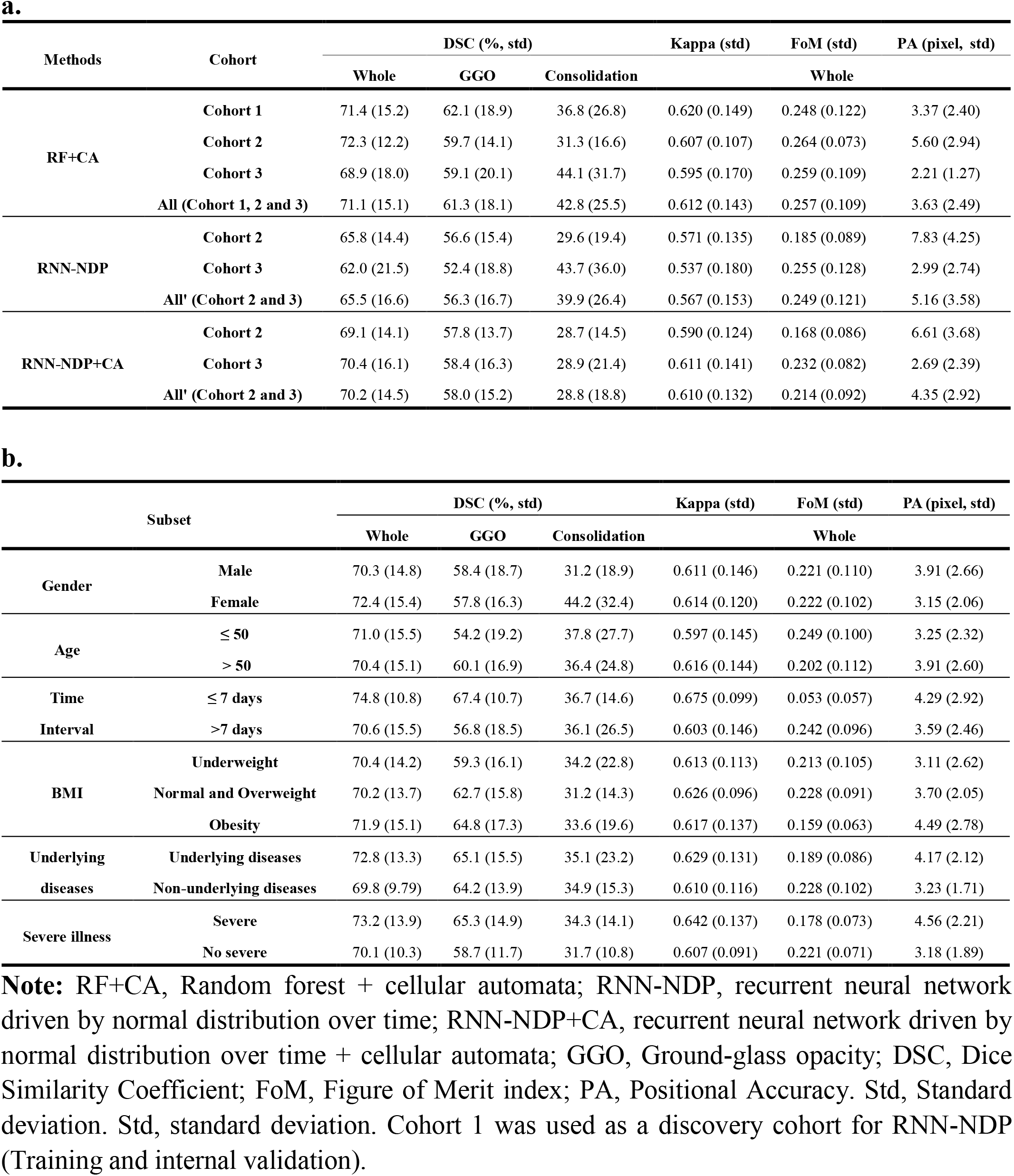
The performance of the forecast system. **a**. Performance comparison of three models: RF+CA, DNN, DNN+CA. **b**. The performance of the RF+CA model in six subsets.

### Construction of the AI System for lesion forecast

Lesions in CT of COVID-19 patients are classified into two major types: ground-glass opacity (GGO) and consolidation. GGO is defined as an area with slightly and homogeneously increased in density that does not obscure underlying vascular markings. The consolidation component is defined as an irregular opaque area that completely blurs the underlying vascular markings.

We developed an AI system, which directly took two-stage CT data as input to perform voxel-level forecast of lesion development. The system consists of four parts: data pre-processing, lung registration, driving factor generation, and lesion development simulation. Firstly, three deep learning models were developed to extract three segmentation masks, i.e. a 3D convolutional neural network (CNN) for segmenting the left and right lung into five lobes on all CT scans, a 2.5D CNN for extracting various tubular adjacent interstitials (TAI) including bronchial bundles, vascular bundles, central lobular stroma, and a semi-automatic method for segmenting the lesion regions. Secondly, all CT scans from the same patient were cropped and then aligned to the first stage CT volume based on the extracted lobe masks, using rigid transformation and non-linear B-spline transformation. Thirdly, we extracted three additional driving factors related to lesion development, including the distance map to the margin of lobe mask, distance map to the center of lesion mask, and distance map to the centerline of TAI. Finally, the lesion regions in the third and subsequent stages were predicted based on the extracted information from previous two stages.

In the abovementioned final part, three voxel-level lesion development forecast models are proposed: recurrent neural network driven by normal distribution over time (RNN-NDT), recurrent neural network driven by normal distribution over time + cellular automaton (RNN-NDT+CA), and random forest classification + cellular automaton (RF+CA). Based on empirical test, the third model (RF+CA) with the best performance was selected to perform the whole study. The workflow of this machine-learning based forecast system is shown in **Figure 1** and **Extended data | Figure 1**.

**Figure 1.**
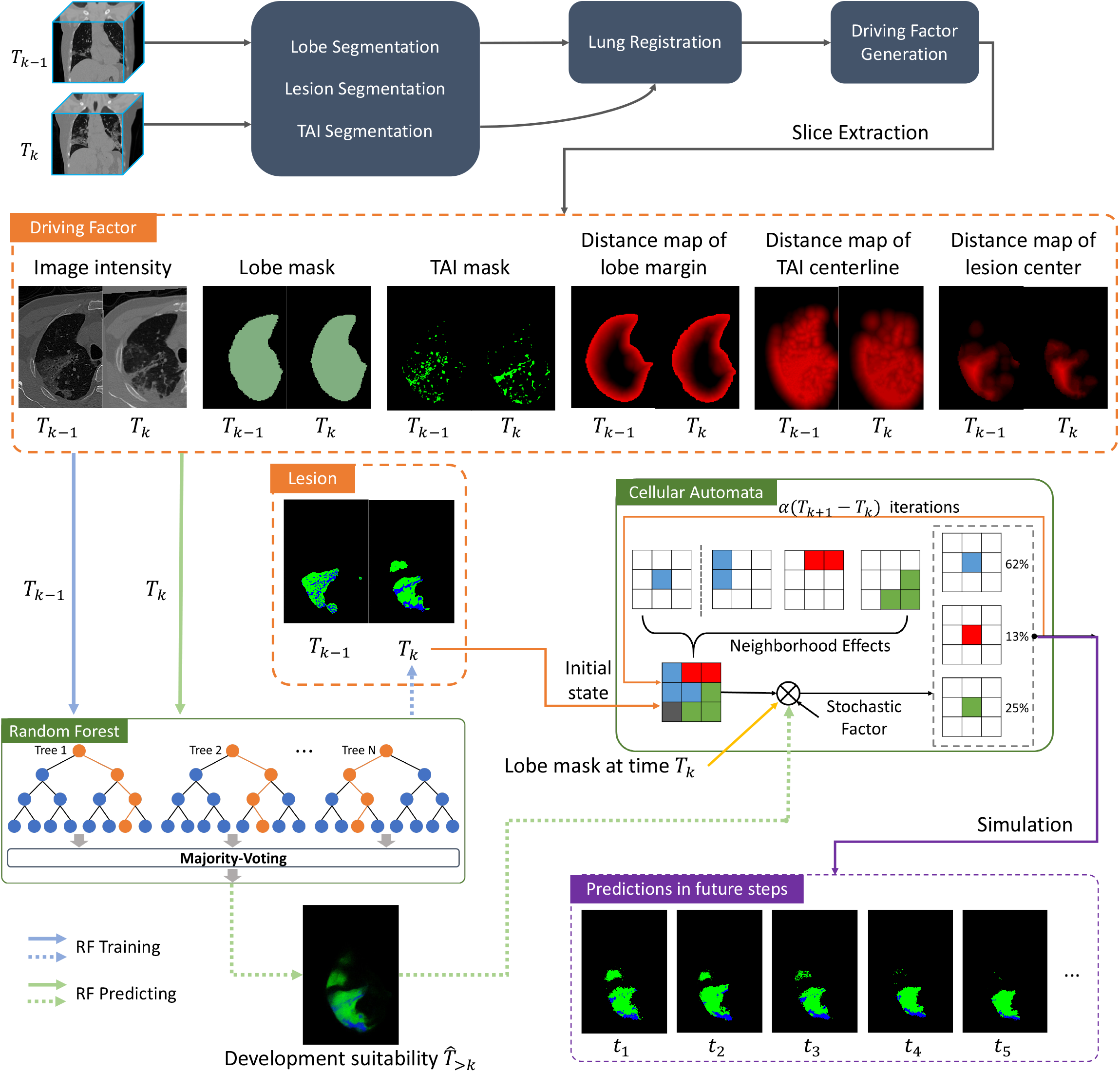
Lesion evolution forecast by random forest and cellular automata (RF+CA). TAI: Tubular adjacent interstitials. Lobe: left and right lung lobes. The green pixels in lesion mask and development suitability image denote GGO and blue ones are consolidation. Note that all the driving factors are generated as volumetric data and followed by the slice extraction operation.

## Performances of forecast system

### Spatio-temporal pattern analysis

To validate the proposed forecast model, we compared the simulation results with the actual lesion distribution in the corresponding stage. **Figures 2** and **3** are comparisons between the simulated results obtained by the proposed model and the actual lesions of two patients. The two patients’ conditions were severe and non-severe, respectively. From the follow-up records, the pulmonary lesions in the severe patient progressed rapidly and this patient was eventually admitted to the intensive care unit (ICU). The non-severe patient was eventually discharged with a better health condition. It can be found that the overall spatial distribution of lesions in the simulation results is close to the real situation in the 3^rd^ stage CT image. The predicted lesion development status in the next 5 and 10 days indicates that multiple lesions are scattered in the lung of the patient in **Figure 2**. Although consolidation area has shrunk, the GGO area has expanded, and more lesions have evolved in the right inferior lobe and the left inferior lobe. The forecast result indicates that the patient would develop into a severer condition. In **Figure 3**, the patient’s lesion area will become more compact in the next 5 and 10 days according to the prediction result. Although consolidation area has expanded, the area of GGO has significantly reduced, and lots of obvious cord signs have been formed. Although several lesion areas have indeed expanded, some straight consolidation cords have appeared on the borders of lung (**Extended data | Figure 2**, **3**), indicating a trend of improvement in the future^21^.

**Figure 2.**
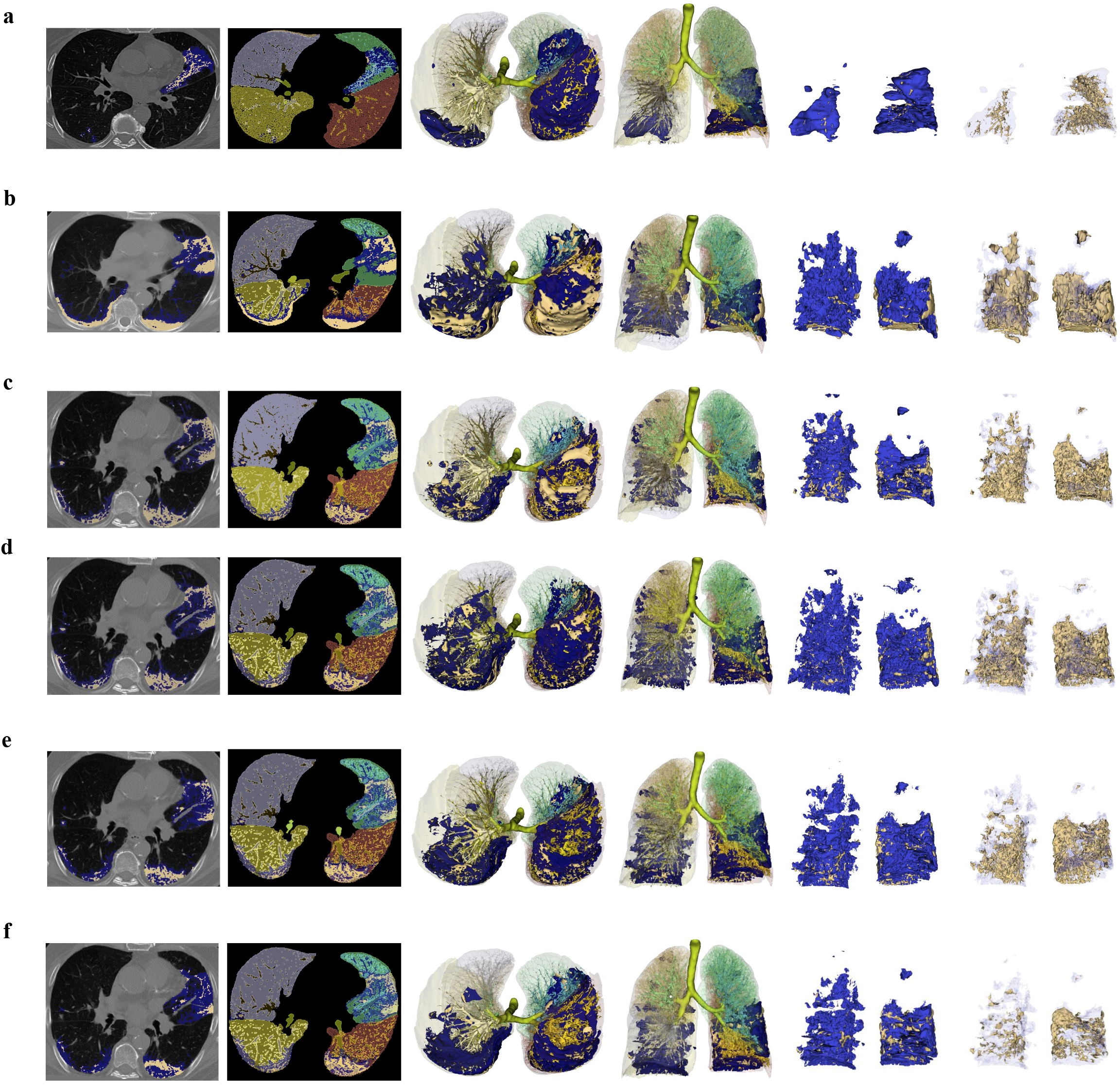
Simulation results of multi-stage evolution of lesion and non-lesion area based on the RF+CA model from a 57-year-old male patient with hypertension. **a** images based on the 1^st^ stage CT image of the patient; **b** images based on the 2^nd^ stage CT image showing the actual lesion evolution; **c, d** images based on the simulated lesion evolution and the 3^rd^ stage of the actual lesion evolution, respectively; **e, f** images based on the predictions of the lesion in the next 5 and 10 days from the 3^rd^ stage of CT according to the trend of the disease. **Column 1**: Lesion location on 2D slicing. **Column 2**: Distribution of lesions in the five lung lobes on 2D slicing. **Column 3**: Distribution of lesions with TAI segmentation in 3-D axial view. **Column 4**: Distribution of lesions with TAI segmentation in 3D coronal view. **Column 5**: 3D reconstruction of the whole lesions. **Column 6**: 3D reconstruction of solid lesions. Dark blue area is GGO and light-yellow area is consolidation.

**Figure 3.**
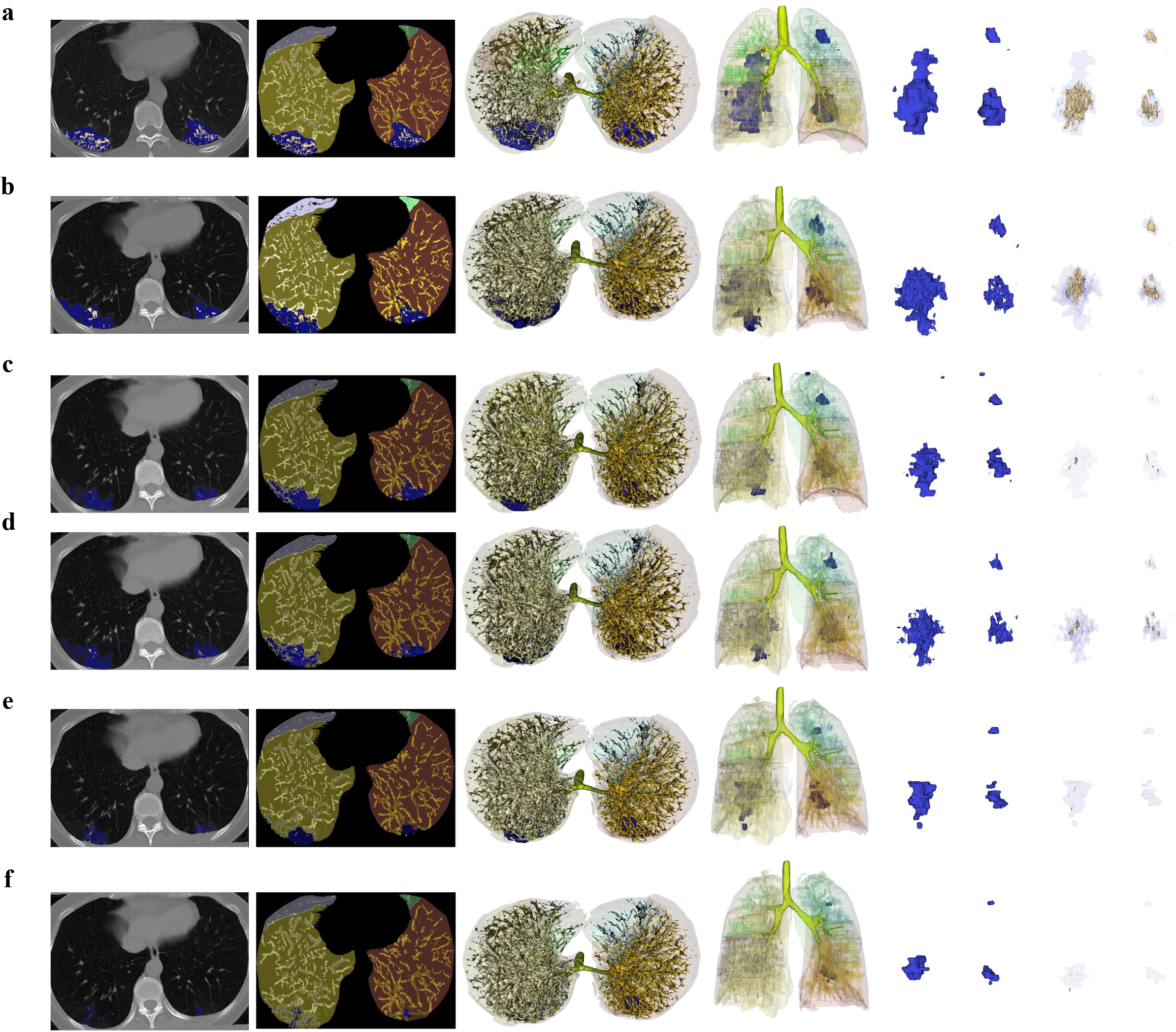
Simulation results of multi-stage evolution of lesion and non-lesion area based on the RF+CA model from a 36-year-old female patient. **a** images based on the 1^st^ stage CT image of the patient; **b** images based on the 2^nd^ stage CT image showing the actual lesion evolution; **c, d** images based on the simulated lesion evolution and the 3^rd^ stage of the actual lesion evolution, respectively; **e, f** images based on the predictions of the lesion in the next 5 and 10 days from the 3^rd^ stage of CT according to the trend of the disease.

**Figure 4a, b** demonstrate the trend curve of whole lesion volume development in five pulmonary lobes of these two patients. The lesion development in different pulmonary lobes of the same patient shows different trends. **Figure 4c** shows the Sankey diagrams visualizing the conversion pattern between the three types of areas (normal area, GGO, and consolidation) in the lung of the severe case. In the progression from the 3^rd^ CT examination to the 5^th^ day after, approximately 12.79% (535.70 cm^3^) of the lungs of this severe patient has undergone intensive lesion evolution. As for the flow-out process, normal area is the category with the largest volume loss (131.39 cm^3^), most of which has evolved into GGO. A large portion of consolidation (85.64 cm^3^) has become normal. A small amount of GGO (21.83 cm^3^) has evolved into consolidation. As for the flow-in process, GGO is the category with the largest volume gain (111.20 cm^3^), indicating that GGO grows at a higher speed and tends to be more stable than other types. In the progression from the 5^th^ to 10^th^ day after the 3^rd^ CT examination, approximately 14.53% of the lung area (608.46 cm^3^) of this patient has undergone intensive lesion evolution. As for the flow-out process, the normal area remains to be the largest volume loss category (147.95 cm^3^). Most of the loss has evolved into GGO (125.01 cm^3^). A small part of consolidation has evolved into normal area (40.09 cm^3^). As for the flow-in process, GGO is the category with the largest gains (136.01 cm^3^), indicating that GGO grows at a higher speed and tends to be more stable than other types.

**Figure 4.**
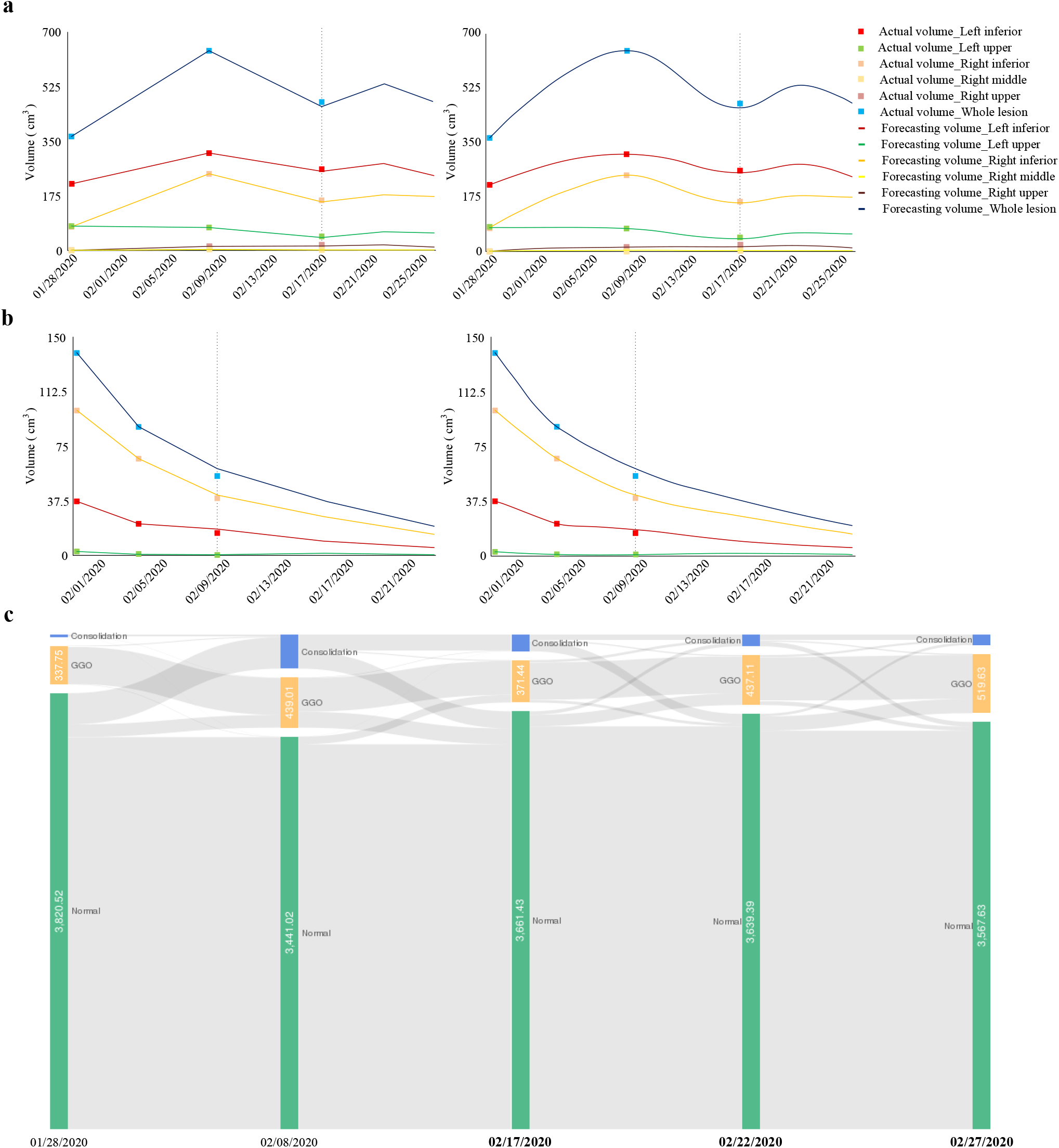
Comparison of simulation and actual trends of lesion development in five pulmonary lobes. According to the development of lesions in the early stages, the forecast system further predicts the trend of lesion development in the next 5 and 10 days from the 3^rd^ CT examination for severe patient and the next 7 and 14 days from the 3rd CT examination for non-severe patient. **a**. The development of recurrent lesions in severe patient; **b**. The gradually vanishing of multiple lesions in non-severe patient; **c**. Sankey diagram for the forecast of development of pulmonary lesions of severe patient. In **a** and **b**, Light colored dots and long dark lines in each group represent actual and simulated trends of the whole lesion development in the same area, respectively. The plots on the right are smoothed version. No obvious lesion was found in the right upper or middle lobe for non-severe patient. In **c**, the transition classes are designated as: Normal, GGO, Consolidation.

### Accuracy of voxel-level lesion development forecast models

Three forecast models, RF+CA, RNN-NDP and RNN-NDP+CA, were developed in this study. To quantitatively evaluate their prediction performance, the evolution of lesions in each patient was compared with actual lesions at corresponding stage point by point. Patients were divided into three cohorts corresponding to three different hospitals and each cohort contained 245, 78 and 104 patients, respectively. The development of the whole lesion region, GGO subregion and consolidation subregion were evaluated, and the resulting confusion matrix is shown in **Table 1a**. DSC of 71.1, Kappa of 0.612, FoM of 0.257, and PA of 3.63 were obtained in the dataset consisting of Cohort 1, 2 and 3 for RF+CA model. DSC of 65.5, Kappa of 0.567, FoM of 0.249, and PA of 5.16 were obtained in the dataset consisting of Cohort 2 and 3 for deep leaning with normal distribution prior model; DSC of 70.2, Kappa of 0.610, FoM of 0.214, and PA of 4.35 were obtained in the dataset consisting of Cohort 2 and 3 for DNN+CA. Due to the small proportion of lesion and scattered distribution of consolidation region in the whole lung, the prediction accuracy of all models including RF+CA is not sufficiently high. However, the RF+CA model has better reliability and higher accuracy than the other two models, especially in terms of Kappa and FoM coefficients, indicating that the RF+CA model is more suitable for the prediction of lesion development. In the following study, we use only the RF+CA model.

### Subset analysis

Most of the patients included in this study recorded clinical information such as BMI and underlying diseases. For an in-depth understanding of the lesion development forecast system and characteristics of different population with COVID-19, we further evaluated the proposed model on six subsets of test cohorts based on gender, age (≤50 and >50 years), the interval between 2^nd^ and 3^rd^ CT examinations (≤7 days and >7 days), BMI (body mass index, underweight [≤18.5], normal weight and overweight [18.5∼29.9], obesity [≥30]), whether suffering from underlying diseases (diabetes, hypertension, cardiovascular disease, cancer, chronic kidney disease, etc.), and whether in a critical condition^22^. **Extended data | Table 1** lists the distribution of patients in the six subsets. **Table 1b** shows DSC, Kappa, FoM, and PA for the six subsets.

### Driving factors of lesion evolution

After training the random forest, the out-of-bag data can be used to measure the importance of each driving factor. As shown in **Figure 5a**, the distance map from the margin of lung region and that from centerlines of various TAI (blood vessels, micro bronchiole, intralobular septum, interlobular septum, etc.) have the greatest impact on the prediction accuracy. Multi-stage images of some patients are shown in **Figure 5b** and **Extended data | Figure 2**,**3**. The “3D+t” animation was further reconstructed and shown in **Lesion development of the patients with COVID-19**.**mov**. The most important driving factor is the distance to the lung margin, which is shown on the left of **Figure 5b** and **Extended data | Figure 2**. The second most important one is the distance to TAI centerline, which is shown on the right of **Figure 5b** and **Extended data | Figure 3**.

**Figure 5.**
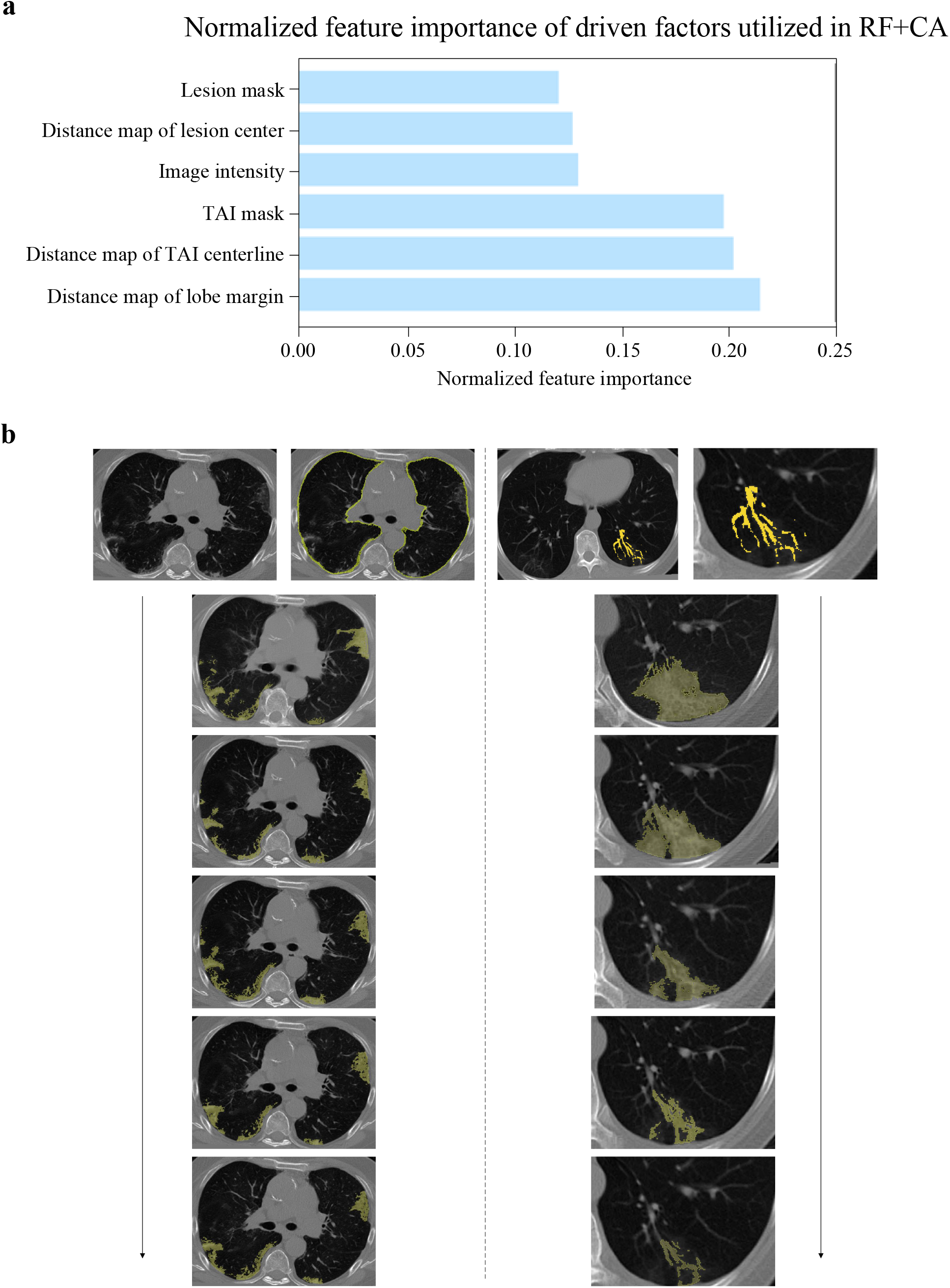
Driving factor of lesion development in patients with COVID-19. **a**. Measurement of importance of each driving factor. TAI denotes various tubular adjacent interstitials, which included bronchial bundles, vascular bundles, central lobular stroma etc. **b**. In the images on the left, lesions are mainly scattered around the peripheral areas of lungs. In the images on the right, lesions grow along TAI.

## Discussion

The prediction of lesion evolution is of major interest to COVID-19 management. The forecast of potential lesion growth could alert clinicians and help in early recognition of development of disease. It is particularly important for COVID-19 since some patients, including those who are young and previously healthy, can go from fine to flailing in the short term^23^. Take as an example the reaction case of Wenliang Li, a young Chinese doctor, called a cytokine storm^24^. According to the forecast of lesion development and clinical manifestations of patients, targeted treatment in advance may greatly reduce the mortality rate.

The development of COVID-19 lesions is a complex nonlinear process. To find the best possible forecast model, three predictive models were established for comparison. The RF+CA model obtained the best accuracy and consistency (Kappa_3^rd^ = 0.612, FoM_3^rd^ = 0.257) in multicenter data. From the perspective of landscape pattern, PA indicates the localization precision of lung lesion. RF+CA yielded a smaller PA (PA_3^rd^ = 3.63), which was superior to the other two methods. These results indicate that RF+CA can accurately mine the law of lesion development under large-scale simulations. It improves single-voxel calculation by focusing on the key spatial driving factors and the influence of cell neighborhood. The lesion development presents different mechanisms among different patients and even different lung lobes, which are ignored in the other two deep learning-based methods. In addition, the random forest method can also measure the importance of each driving factor according to its contribution to the prediction, which helps to explain the role of each driving factor in the lesion development. Parallel construction of random forest can greatly reduce its training time. Therefore, the CA model based on random forest has the advantages of high accuracy, fast training, and good interpretability. In the follow-up sections of this study, the proposed method refers to RF+CA.

Based on the proposed forecast model, we explored driving factors that affect the development of the lesion anatomically, and further carried out image description and pathophysiological explanation for specific patients. The proposed lesion development forecast system is of prompt value for disease development evaluation and prognosis, as well as for the pathogenesis of COVID-19. From the perspective of imaging alone, by simulating and forecasting more multi-stage CT images of COVID-19 patients, dynamic analysis of radiological characteristics in the long course of the disease can be performed in a predictable manner at the early stage of the disease, and major imaging markers belonging to phenotypic characteristics that distinguish mild and severe diseases can be identified (One of the applications of this forecast system for lesion development system. See the section on radiomics analysis of the long course of disease in supplementary materials for details.).

In this study, we conclude that the distance from the lung margin (**Figure 5a, b**) is the major driving factor for the transformation of non-lesion regions into lesion regions. Because the peripheral subpleural pulmonary lobules are better developed, blood flow and lymphatic are abundant, and the corresponding lobular interstitial inflammatory reaction is more obvious, the distribution pattern is mainly subpleural^25-27^. The unique pattern of growth of COVID-19 lesions is parallel to pleural spread, that is, the stripe pattern is parallel to the pleura (**Extended data | Figure 2, 3**). The growth mechanism is presumed to be: Severe acute respiratory syndrome 2 (SARS CoV-2) virus invades lobular interstitium. When the peri-lobular stroma is mainly invaded, that is, the peri-alveolar stroma, this part of the lymphatic drainage direction is subpleural and interlobular septum^25^. The spread is also mainly around, diffusing to the pleura side and both side of the interlobular space. Because the distal end is restricted by the pleura, the lesion growth can only be close to the pleura, spreading to both sides along the edge of the interlobular septum of mesh structure. The mutual fusion of the subpleural lesions causes the long axis of the lesion to be parallel to the pleura^28^. With the continuation of the disease, the lesions show a spreading trend from peripheral to centra. Two growth modes of lobular core interstitial and subpleural fuse together and gradually spread to most areas of the lung lobe and even diffuse to both lungs (“white lung”)^29^. Another major factor is the distance to various interstitium (TAI centerline) (**Figure 5a, b**). It is intuitive that the virus first invades the bronchiole, causing bronchiolitis and peripheral inflammation. Then the virus spreads along various interstitials, and finally spreads to the lung parenchyma^30^. The thickening of the interlobular interstitial cord sign and fine mesh sign (crazy paving appearance) confirm that the growth of the virus mainly involves the interstitium of the lobules^31^. The interstitium in the lung includes the axial interstitium and the surrounding interstitium. The axial stroma includes bronchial bundles, vascular bundles, lymphatic vascular bundles, and central lobular stroma. The surrounding stroma includes the subpleural interlobular septum and the intralobular interstitium^28,32^.

We divided the lesions into two categories according to the CT values: i. Ground glass opacity (GGO). In this type of lesion, local lung tissue has slightly increased density, but bronchial and blood vessels can still be clearly displayed; ii. consolidation. In this type of lesion, local lung tissue has increased density, and bronchial and vascular are unclear. In addition to the focal patches and mass signs, the highlights of fibrous bands, mesh signs, and subpleural linear bands are included in the consolidation range. The radiologic findings were evaluated by thresholding on CT values^16^. GGO and consolidation regions were determined by ranges of −700∼-200 Hounsfield units (HU) and −200∼60 HU, respectively. Furthermore, when GGO intertwined with consolidation, the two were separated according to the sudden change in voxel’s CT level. The large area with low CT intensity was classified as GGO, while the other with relatively high CT values was classified as consolidation. In general, the lesion area varies in texture, size and location; The boundary of GGO generally has low gray contrast and blurred appearance. The consolidation area is relatively small, scattered and irregular. Automatic segmentation of these lesions is a challenging task. According to several published studies on segmentation of COVID-19 lesions, the segmentation accuracies are as follows: Ground-Glass Opacity has a DSC less than 65%, and Consolidation has a DSC less than 46%^33-35^. In the proposed forecast model, GGO has a DSC of 61.3%, and Consolidation has a DSC of 42.8%. Compared with the performance of lesion segmentation (which is a much easier task), the performance of our proposed forecast model is reasonable.

In this study, some cases showed a co-occurrence of progression and organized repair of local lesions, that is, partial lesions improvement accompanied by some lesions aggravation or breeding of new lesions. In these cases of recurrent growth and evolution of multiple lesions, the proposed model can also achieve robust performance. After the comparative analysis of multi-stage images of the patients, we learned that the new lesions would replicate the growth process of earlier lesions, but only a delayed inflammatory response in some lung areas, rather than the aggravation or re-infection, as shown in **Figure 2, 4a** and **Lesion development of the patients with COVID-19**.**mov** in supplementary materials. Therefore, if a few lesions in the previous stages show a trend of improvement or deterioration, lesions in the subsequent stages will also develop in a step-by-step manner. Due to this development pattern, our proposed model is able to excavate the correlation between multiple lesions and various driving factors.

Our proposed model was tested in various subsets and acceptable results were obtained. As shown in **Table 1b**, Kappa coefficient is greater than 0.60 in most of the 6 subsets, with a satisfactory agreement. The FoM values in different subsets have significant heterogeneity. Patients with advanced age, underlying diseases, obesity, and severe illness tend to have lower FoM values. This is due to the rapid and complex development of lung lesions in these patients, which potentially reflects some of their individual physiological functions. For instance, patients with severe obesity (BMI≥35) may suffer from dyspnea due to the fat under the diaphragm. Fat produces a considerable amount of pro-inflammatory molecules called cytokines, i.e., an immune battle in the human body producing low-level background inflammation^36^, which is another important risk factor. The development of lesions is an organic and adaptive mechanism that affected by the characteristics of individuals. The changes of lesions are not only related to spatial variables, but also related to patients’ basic clinical characteristics (age, gender, Body Mass Index), whether there is underlying disease, treatment methods, and dynamic changes in laboratory test indicators. These factors have great complexity and randomness. Therefore, the proposed RF+CA model still cannot fully consider the actual situation despite its superior performance over other models.

In conclusion, voxel-level forecast model for the development of lesions is of great significance for personalized treatment of COVID-19 patients and the matching and coordination of limited medical resources in the future. We made a preliminary exploration to this challenging topic. Due to the limited patient data and the large interval between multi-stage CT examinations, the predictive ability of the proposed system still has room for improvement. We believe that the performance will improve if more cases are collected and analyzed. In addition, the prediction method can also be extended to the prediction of lesions development for other diseases, such as acute stroke.

## Methods

### Data Collection

We collected CT volumes from three different centers in Wuhan,which are Wuhan Union Hospital, Western Campus of Wuhan Union Hospital, and Jianghan Mobile Cabin Hospital. 1337 standard 3D CT volumes of 417 subjects diagnosed as COVID-19 were selected. All CT scans were performed with patients in supine position, using one of the following scanners: SOMATOM Perspective, SOMATOM Spirit, or SOMATOM Definition AS+ (Siemens^®^ Healthineers, Forchheim, Germany). Scanning was performed from the level of the upper chest entrance to the lower angle of the costal diaphragm, using the following parameters: detector collimation width 64 × 0.6 mm, 128 × 0.6 mm, 64 × 0.6 mm, 64 × 0.6 mm; tube voltage 120 kv. The tube current was adjusted by the automatic exposure control system (Care dose 4D; Siemens Healthineers). The slice thicknesses of the reconstructed image were 1.5 mm and 1 mm, and the interval were 1.5 mm and 1 mm. In all cases, the upper limit of the interval between adjacent CT scans was 15 days, the average value was approximately 7 days. To utilize all data efficiently, all cases with more than 3 stages were split into sub-groups with 3 consecutive stages using sliding window method. Finally, 503 sub-groups are collected as the experimental dataset in this study.

### Image annotation

Three types of masks are required considering the characteristic of lesion development, including the whole lung region, various TAI and lesions caused by COVID-19. For simplicity, two categories of lung masks and two categories of lesions were determined, while the different TAI categories were regarded as the same class. Specifically, lung masks were divided into left and right lobes, while the lesions caused by COVID-19 were distinguished as GGO and consolidation, respectively.

For lung and lesion mask, annotation procedure was applied in an automatic way: a vanilla 3D U-Net (**Figure S1a**, see supplementary methods for details) based on 60 manually annotated scans was trained to generate segmentation masks. Since lung segmentation was easy and achieved acceptable performance, there was no further modification performed. Due to the complicated background and various appearance of lesion, a refinement was performed after automatic segmentation by two well-trained experts (the two board-certified radiologists have 12 and 21 years of experience, respectively) using a self-developed multi-stage simultaneous segmentation toolbox (https://github.com/weixr18/OCD_Slicer_Plugin/tree/release-amd64/OpenCOVID_Detecter/CT_Annotate) based on 3D Slicer (https://www.slicer.org/). After their separate annotation, any differences were resolved through discussion and consensus. For TAI mask, a 2.5D segmentation framework, containing three vanilla 2D U-Nets (**Figure S1 b**, see supplementary methods for details), was designed to separate the tubular structure from complicated background, and trained on 49 scans that were manually annotated using Mimics (Materialise, Leuven, Belgium).

### Data pre-processing

The HU value of CT volumes ranges from −2048 to 3071. For a clear view of lungs and tissues inside lungs, we truncated a window of [−1200,700], and normalized the value to float value as standard normal distribution.

Firstly, a lung segmentation network, i.e. vanilla 3D U-Net (The network structure is shown in **Figure S1a** in Supplementary methods), was trained on a dataset containing 60 chest CT scans. Then all volumes in our dataset were processed to generate the masks of left and right lung with five lobes. Similarly, lesion and TAI mask were generated using the abovementioned segmentation framework, and leaks outside the lung region were removed. The segmented TAI masks are shown in **Figure 2, 3** in **Result** section.

Then a cropping operation was performed to exclude complicated background. Note that the lung segmentation results were split into left and right lung lobe groups, and thus all volumes were cropped to contain the left or right lung lobe only.

Since our proposed algorithm takes 2D images as input, all volumes are supposed to be sliced. Registration, consequently, is needed before doing such slicing operation, to keep consistence at voxel level between different stages. Specifically, a registration was applied to align the volumes for the same patient^37^ to the same coordinate system (extracted from the first stage of CT). Considering the characteristics of lung region and the fact that only the regions inside lungs were what we concern, we utilized an affine transformation first to initialize the alignment, followed by a B-spline transformation to refine^38^, based on the extracted lobe masks. All other segmentation masks extracted from the same CT volume were then transformed to the same coordinate based on the estimated transformation parameters. All these parameter estimation and transformation procedure were achieved using *SimpleElastix* in Python^39^.

Based on the cropped and aligned 3D volumes, some driving factors, used in our RF+CA method later, were generated automatically. Driving factors are defined based on the distribution and pathology of abnormal regions, e.g., expanding through the tubular-system such as vessels and airways, spreading below the subpleural. The driving factors consist of 6 categories, including original image intensity, lobe mask, TAI mask, distance map of each pixel to the margin of lobe mask, distance map to the center of lesion region, and distance map to the centerline of various TAI.

Note that due to different lesion categories sharing similar characteristics, the distance map to the center of lesion mask was calculated based on the whole lesion regions yet ignoring the specific types. To extract the distance map of TAI, the centerlines were extracted first using the morphological thinning algorithm, and followed by the Euclidean distance transformation algorithm from *Scipy* in Python^40^. The other two distance maps were generated similarly.

Finally, all these volumes were sliced. Slices in later stages were ignored for brevity, if the corresponding first stage contained no obvious lesions, unless the slices had considerable disease (the lesion region larger than 30 pixels). Different stages from the same patient were grouped for further analysis. Consequently, 35018 groups of multi-stage sequential images and their corresponding lesion masks along with driving factors were selected for further analysis. Note that each group contained 3 stages only.

### Estimate suitability using random forest algorithm

Development suitability, namely the potential changing energy of each lesion categories, was estimated by applying random forest (RF) algorithm^41^ on slices of original images and corresponding driving factors from current stage.

The RF model took each pixel along with its 15 × 15 surrounding neighbors from current stage as input. It was then supervised by the annotated lesion category of center pixel from the next stage. The first two stages were used for training and the last stage was used for estimation and evaluation. Due to the class imbalance, which means background region dominated the image, balanced-subsampling and bootstrapping were utilized to explore the hard samples, and 100 independent decision trees were used to increase estimation accuracy and generalization ability. Based on the estimated development suitability, the development probability for next stage was calculated, which was utilized in cellular automata (CA) simulation later.

The most important step in this algorithm is to optimize different model parameters using the first two stages for different time, patients, and even lobes (i.e., left and right lung). It is intuitive that different model parameters need to be estimated since the development of disease in different patients varies. Besides, the relation of disease development between left and right lung lobe is not obvious in most cases, e.g., the lesion region is growing in left lung lobe, while the size is reducing in another lung lobe (in fact, considering the presence of fissure, the disease development progression varies in five lobes). Furthermore, the lesion progression speed and tendency would not be identical within the same lung lobe. Therefore, the RF model was finetuned when there were new stages incorporated as training samples to estimate lesion regions at later stages.

### Evaluation metric

To evaluate the performance of the aforementioned algorithm, we considered the classic metrics used in object segmentation and land use and cover change (LUCC) task, respectively. DSC^42^ is widely used in object segmentation task to evaluate the similarity between prediction and annotation mask:

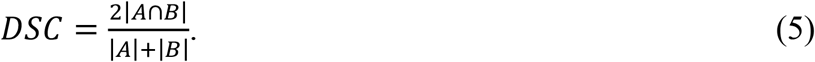

Here *A* and *B* represent the set of ground-truth and the prediction, respectively. |·| denotes the number of pixels associated with the certain class label, e.g. the number of pixels predicted or annotated as GGO. When the prediction is similar to annotation in pixel level, the metric DSC is close to 1, otherwise 0.

In LUCC tasks, Kappa coefficient and FoM are mostly adopted to verify model accuracy^43,44^. Kappa coefficient is a statistical measure of the reliability or consistency of internal raters, and defined as follows:

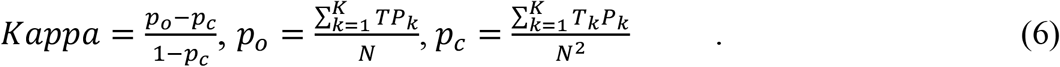

Here, *K* and *N* represent the number of classes and the total number of samples, respectively. *TPk, Tk* and *Pk* represent the number of pixels correctly predicted as class *k* (correct prediction), the number of pixels annotated as class *k* (ground-truth) and the number of pixels predicted as class *k* (prediction), respectively. In other words, *p*_*o*_ represents the total observed accuracy and *p*_*c*_ represents the chance agreement. The strength interpretation of Kappa coefficient is: slight [0.01-0.20]; fair [0.21-0.40]; moderate [0.41-0.60]; substantial [0.61-0.80]; nearly perfect [0.81-1.00].

FoM is a quantity used to measure the prediction performance. The formula is defined as follows:

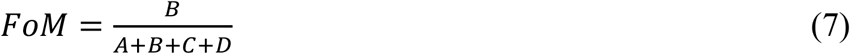

where *A* is the error zone caused by the prediction of observed change as persistence; *B* is the correct zone caused by the prediction of observed change as change; *C* is the error zone caused by the prediction of observed change as wrong gaining category; *D* is the error zone caused by the prediction of observed persistence as change.

Additionally, to evaluate the location accuracy of predicted area, we proposed a metric named positional accuracy (PA) to measure the offset between predicted lesion region and ground-truth, in pixel level, which is defined as followed,

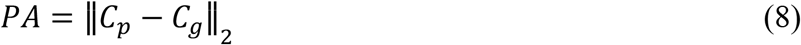

where *C*_*p*_ and *C*_*g*_ are the equivalent mass center of prediction and annotation mask, respectively.

### Cellular automata simulation

In addition to the development suitability, the lesion development is influenced by other factors, such as neighborhood effects, constraint restriction, and stochastic randomness.

Neighborhood effects *Ω* play a vital role in CA algorithm. In the CA model, there are local interactions between cells, and the state(diseased or not)of a cell at the next moment is determined by its neighbors. e.g., a normal cell has an extremely high probability to change to GGO in the next stage, when the surrounding cells are simulated as GGO at current stage. The most frequently employed neighborhood models are von Neymann, Moore and extended square neighborhood. To avoid disparate effects, we used Moore neighborhood^45^ in this study.

Constraint factor *P*_*c*_ is a limitation to prevent lesions from presenting outside lung mask. The undevelopable area is determined in advance, i.e. the cells outside of lung region are restricted from lesion development.

Stochastic factor *P*_*r*_, intuitively, is introduced to increase randomness and thus generalization ability. The evolution of lesions is not only affected by various deterministic factors including the development suitability, but also random factors such as whether the patient suffers from basic diseases, psychological state, treatment mode etc.

Considering the above four parts, the probability that a single cell will be transformed at time *t +* 1 is defined as:

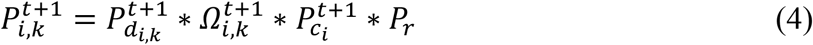

Here 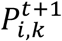 is the probability that cell *i* converts to class *k* at time *t +* 1. 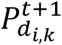 is the development suitability of cell *i* belonging to class *k* at time *t +* 1. Therefore, 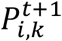 was fed into CA model to simulate development of infectious area.

Combing the calculated development suitability, neighborhood effects, constraint factors and stochastic factors, the probability of transformation for every single cell, i.e. pixel, was determined. Then the simulation using CA algorithm^46 47^ was applied by setting the simulation steps to half of the time interval between the current and next stage.

In summary, the RF+CA algorithm is divided into following steps: (1) Construct a set of data (driving factors) related to the development of infectious area. (2) The RF model is trained, using original images and driving factors from the first stage, to predict lesion potential (development suitability in CA algorithm) for the second stage. (3) The estimated RF model is utilized to predict development suitability for the third stage based on the existing image and driving factors from the second stage. (4) The development suitability, neighborhood effects, constraint factors and stochastic factors are combined to simulate the lesion development using CA algorithm. (5) Considering the time interval and disease evolution speed based on the previous data, the final probability of whether some regions of lungs will develop into lesion or not in the future is predicted. The workflow of RF+CA model is shown in **Figure 1**.

## Model to model comparison

### Deep leaning with normal distribution prior

During the manual annotation of lesion mask, we noted that the development of most lesions followed a gaussian-like distribution^48,49^ of time *T* (See **Figure S2** and Supplementary methods for details). Therefore, it is a reliable hypothesis that the lesion caused by COVID-19 follows the same distribution family with different parameters in different areas (The growth pattern of multiple lesions in the same patient is consistent. See the Discussion section of the main text for details).

Therefore, a 3D CNN was trained to estimate the distribution parameters for each pixel given original image slices from more than two stages. To introduce the time relevant information, *Gated Recurrent Unit* (GRU)^50^, a variant of convolutional Long Short-Term Memory (conv-LSTM), was utilized to enhance time interval related information flow. The outputs of decoder were parameters of the predetermined gaussian distribution, *μ* and *σ*^2^. For stability, the expectation *μ* was set as the offset from current position to the peak, and log*σ*^2^ was estimated instead. In addition, considering the complicated mechanism of lesion development, we used sigmoid function, an extra asymmetric function, to control the development speed before and after the peak moment. Consequently, the lesion at any moment, theoretically, could be predicted given these distribution parameters estimated approximately. However, limited by the data used for estimation, the precision and consistency decreased with increasing time interval. The workflow is also shown in **Figure S2** of Supplementary methods.

### Deep learning-based CA

We used the deep learning model described in the previous section to generate development suitability. Different from RF model, which was trained only on one group of slices at each stage, this model was trained on a subset of extracted slices. Similarly, the output was regarded as development suitability *P*_*d*_ used in CA algorithm. It was combined with neighborhood effects *Ω*, constraint factor *P*_*c*_ and stochastic factor *P*_*r*_ to determine the transformation probability for each pixel. The overall workflow is shown in **Figure S3** of Supplementary methods.

## Data Availability

The datasets from Wuhan Union Hospital, Western Campus of Wuhan Union Hospital, and Jianghan Mobile Cabin Hospital were used under the license of the current study and are not publicly available.

---

**Extended data | Table 1.**
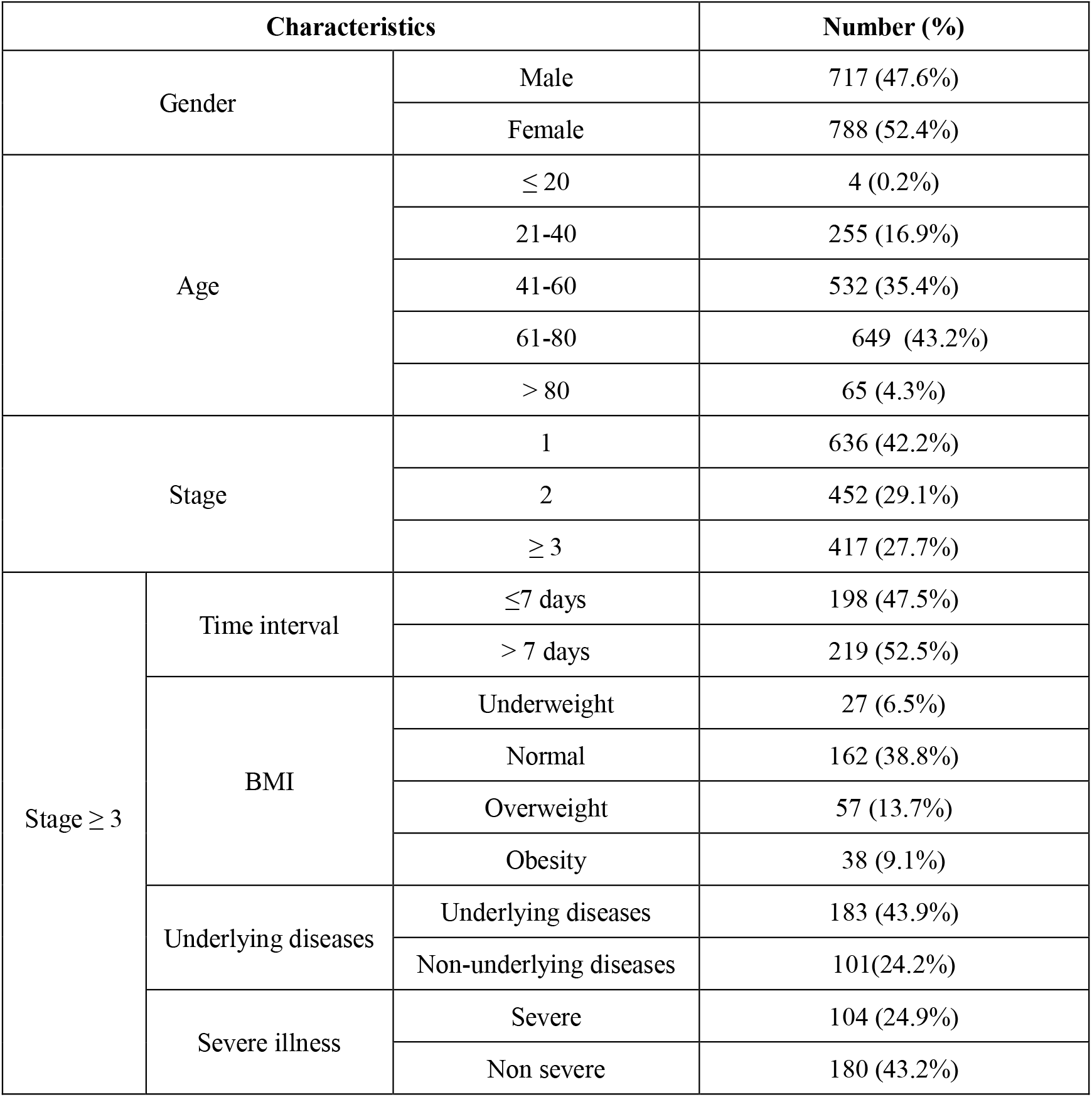
Baseline clinical characteristics of patients in the study.

**Extended data | Figure 1.**
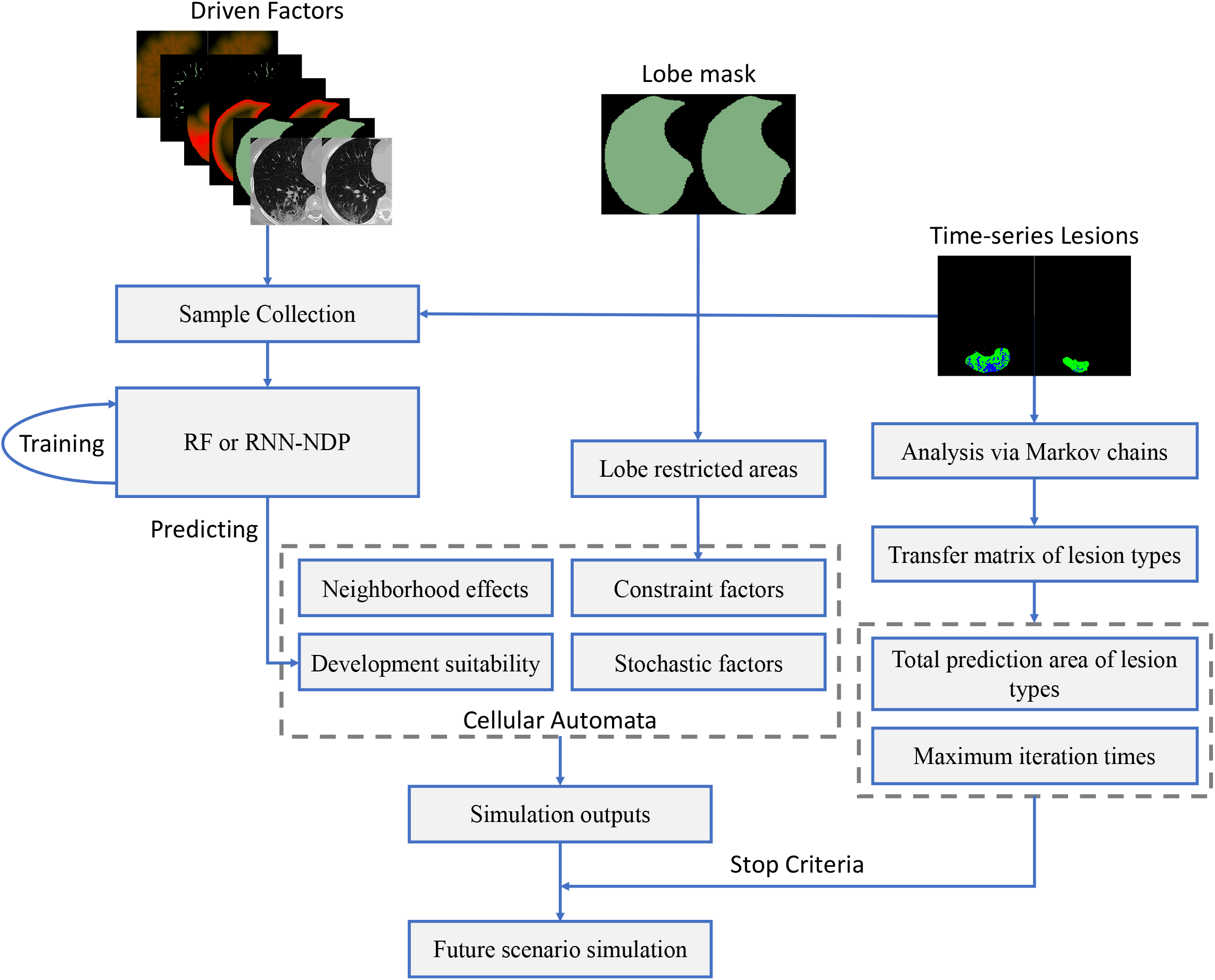
The flowchart of lesion evolution simulation via the CA based forecasting system.

**Extended data | Figure 2.**
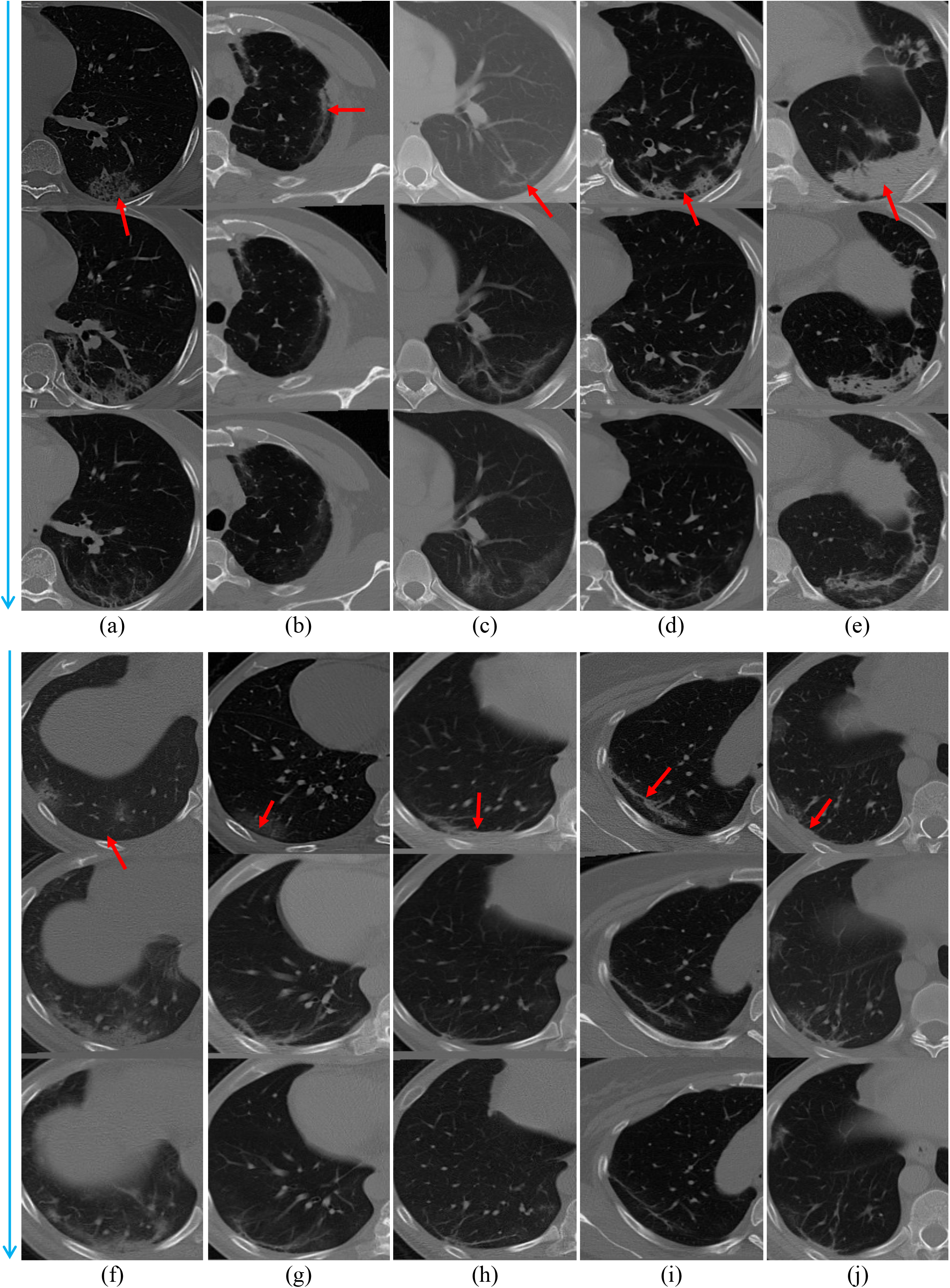
Multi-stage samples where lesion develops below the subpleural and along some vessels. The red arrows point to the lesions. Each case contains 3 stages, forward from top to bottom (blue arrow).

**Extended data | Figure 3.**
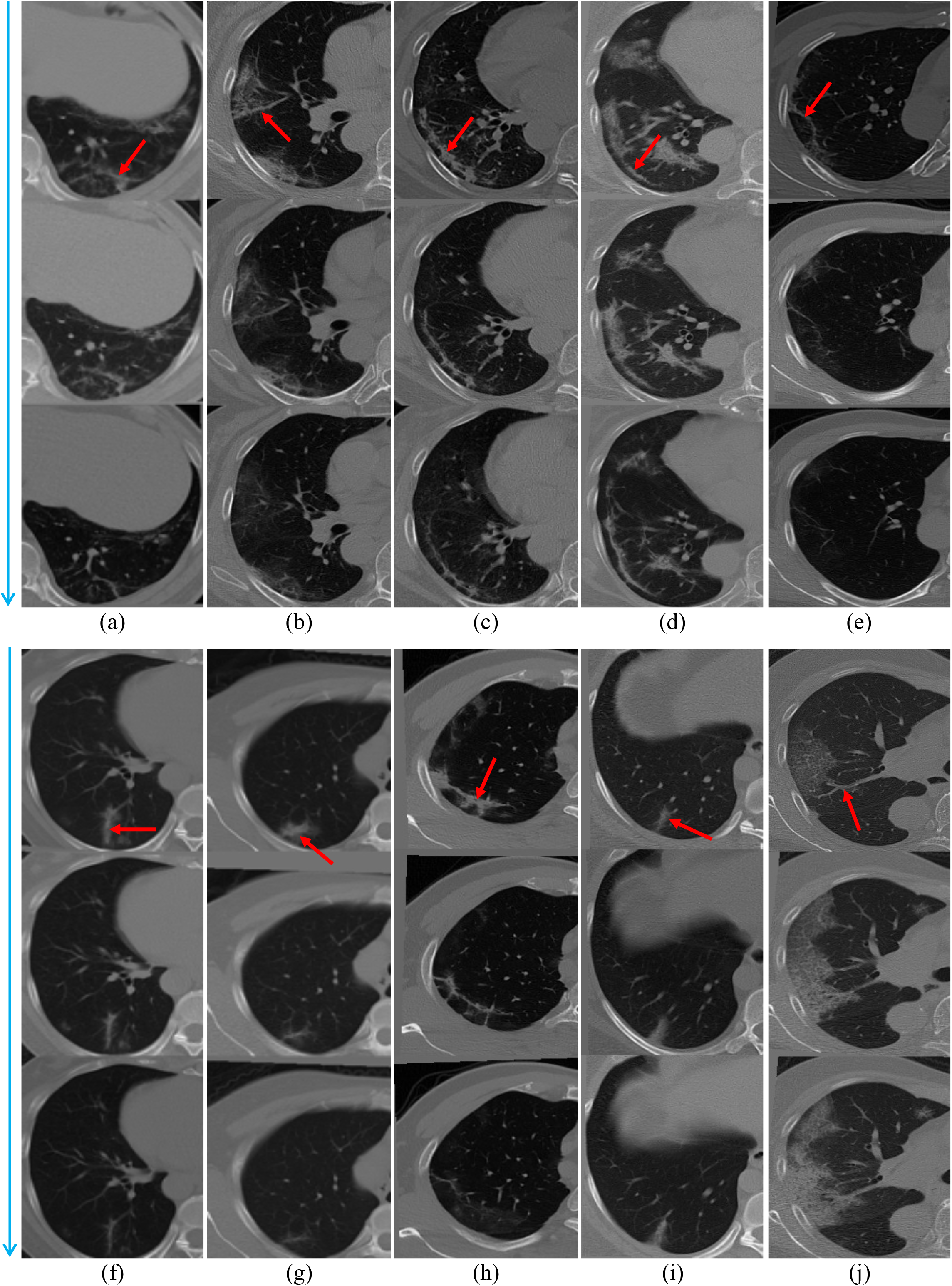
Multi-stage samples where lesion develops along the tubular adjacent interstitials (TAI). The red arrows point to the lesions. Each case contains 3 stages, forward from top to bottom (blue arrow).

