## Supplemental Information for "Voxel-level forecast system for lesion development in patients with COVID-19"

---

---

Cheng Jin, Yongjie Duan, Yukun Cao, Jinyang Yu, Zhanwei Xu, Weixiang Chen, Xiaoyu Han, Jia Liu, Jie Zhou, Heshui Shi, Jianjiang Feng 

#### Supplementary Methods

### Data annotation

Lung extraction was applied first to reduce search space of lesions and eliminate complicated noise outside lung mask. A lung segmentation model based on vanilla U-Net (**Figure S1 a, b**) was trained on an extra dataset whose annotations are labeled manually by two well-trained experts. In this study, we segmented left lung and right lung separately. All CT volumes were then segmented and cropped according to the extracted lung mask.

Due to the complicated background and various appearance of lesions, a similar U-Net was pre-trained on a manually annotated dataset to generate coarse masks first. A refinement was performed by two well-trained experts (The two board-certified radiologists have 12 and 21 years of experience, respectively). After their separate assessment, any differences are resolved through discussion and consensus.

For tubular adjacent interstitials (TAI) mask, a 2.5D segmentation framework, containing three vanilla 2D U-Nets, was designed to separate the approximate tubular structures (bronchial bundles, vascular bundles, lymphatic vascular bundles, and central lobular stroma et al.) from complicated background first. It was then trained on data that were manually annotated using the Mimics software and a self-developed multi-stage simultaneous segmentation tool based on 3-D Slicer. Based on this model, all volumes are processed to extract TAI.

### Data pre-processing

To explore the development potential of lesions, volumes from the same patient have to be aligned according to the first stage. We found that all lesions caused by COVID-19 present below the pleura or around the lung margin. Besides, the lung region is large enough to describe holistic shape and location information of lung regions. Therefore, for simplicity, only the extracted lung masks were utilized to optimize affine and B-spline transformation parameters for every image from the same patient.

CT volumes were sliced after the alignment. Slices in later stages would be ignored, if the corresponding first stage contained no or barely infectious tissue, since the mainly concerned is the development of COVID-infectious area in this study. A total of 35018 sets of multi-stage sequential images were selected, from which 19016 sets (Cohort 1) were used for lesion development training and internal validation, 6858 sets (Cohort 2) were used for independent validation, 9144 sets (Cohort 3) were used for another independent validation.

### RF+CA model

Given one set of multi-stage slices, the development potential is calculated according to the transformation computed from previous two stages. Then the development model is utilized to predict states transformation based on the second slice, consequently an estimation of the last stage slice. Our proposed method is integrated with random forest (RF) and cellular automata (CA) model. CA simulation method is widely applied in land-use and land-cover change (LUCC) problems[1-3]. In this study, it is utilized as the development model to encode the spontaneity and self-organizing feature. We adopt RF method to calculate the lesion development suitability, which acts as an input of CA model to simulate the future lesion region.

Apart from the development suitability generated by RF algorithm, another three factors also play vital roles in CA simulation, i.e. neighborhood effects, constraint restriction and random factors.

Proximity is one of the essential factors in lesion evolution model. Previous studies in LUCC task have shown the effectiveness of CA algorithm in modeling neighborhood configuration effects[1]. Specifically, the neighborhood effects describe the influence from surrounding cells, which means the center cells will change to lesion if most of the surrounding cells were diseased as well. The calculation of the neighborhood function is as follows:

$$\Omega_{ij,k}^{t+1} = \sum_{ij} w_{ij}\, \mathbb{I}\left(S_{ij}^{t} = Lesion\ k\right) \tag{1}$$

Here, $\Omega_{ij,k}^{t+1}$ represents the neighborhood influence of lesion class $k$ at position $ij$, brought by its surrounding $M \times M$ neighbors, at time $t$+1 and $M$ is set as 5 in this study; $w_{ij}$ denotes weight defined by the distance to the center from surrounding neighbors, which means the closer the neighbor is to the center cell, the more important it is; $\mathbb{I}(\cdot)$ represents indicator function; $S_{ij}^{t}$ is the current state of the cell, i.e. the lesion category.

The constraint conditions can be expressed as follows:

$$p_{c_i}^{t+1} = \mathrm{con}(S_i^t) = \begin{cases} 0 & \text{The cell is not allowed to develop into lesions} \\ 1 & \text{The cell is allowed to develop into lesions} \end{cases} \tag{2}$$

Here, $\mathrm{con}(S_i^t)$ judges whether the cell at position $i$ at time t+1 can develop into lesion, and $S_{ij}^{t}$ is the current state value of the cell. The undevelopable area is determined in advance, that is, the cells outside the lung area are restricted from development. In the restricted development area of $S_{ij}^{t}$, all the values are 0.

To reflect these stochastic processes, random factors are introduced in this model:

$$P_r = 1 + (-ln r)^{a} \tag{3}$$

Here, $P_r$ is the part of the random factor, $r$ is the random number within the range of (0,1), and $a$ is the parameter controlling the influence size of the random variable (integer ranging from 1 to 10). In this study, $a$ is set to 2.

The transformation probability $P$ of each cell, i.e. each pixel in a slice image, is composed of four parts: development suitability $P_d$, neighborhood effects $\Omega$, constraint factor $P_c$ and stochastic factor $P_r$. The development suitability $P_d$ is estimated by aforementioned RF model.

Considering the above four parts, the probability that a single cell will be transformed at time $t$ is defined as:

$$P_{i,k}^{t+1} = P_{d_{i,k}}^{t+1} * \Omega_{i,k}^{t+1} * P_{c_i}^{t+1} * P_r \quad (4)$$

Here $P_{i,k}^{t+1}$ is the probability that cell $i$ converts to class $k$ at time $t$+1. $P_{d_{i,k}}^{t+1}$ is the development suitability of cell $i$ belonging to class $k$ at time $t$+1. Therefore, $P_{i,k}^{t+1}$ is fed into CA model to simulate development of infectious area.

The simulation iteration times is set as the half of actual days between the last two stages. The overall flowchart is shown in **Figure 1**.

### RNN-NDP model

The performance of lesion probability prediction using simple recurrent neural network (RNN) model is not ideal, since the intervals between multi-stage CT scans of different patients are not equal. Therefore, we proposed a recurrent neural network driven by normal distribution over time. Strong external treatment measures such as surgery are not generally taken for routine treatment of pneumonia. Therefore, the evolution of lesion depends on its own growth mechanism and current surroundings, that is, the probability of non-lesion voxels evolving into lesions is directly related to the number of lesion voxels in its neighborhood. The development of most lesions follows the gaussian-like distribution of time T and spatial S dimension [4,5] except the interference of lung cracks, thick air pipes, etc..

This deep learning network mines spatio-temporal information by combining parallel and serial components, which consists of Encoder, GRU, Decoder and other parts, as shown in **Figure S2 a, b**. The network attempts to learn the two parameters $\mu$ and $\sigma^2$ of the Gaussian distribution describing whether the voxel will evolve into lesion, instead of directly outputting the probability that the voxel belongs to the lesion. GRU is utilized to incorporate time interval information and gaussian distribution prior is assumed to simulate the development of lesion caused by COVID-19.

In this study, we used Cohort 1 (235 cases), as the discovery cohort for model training, in which one fourth of the cases were used as the training set and the rest were used as the internal validation set. In addition, Cohort 2 (78 cases), Cohort 3 (104 cases) were used as two independent validation cohorts. During the training phase，the batch size was set to 16. The learning rate was set to 0.001 and learning rate decay was 0.1 when the validation performance does not decrease over 10 epochs. We used instance normalization instead of traditional batch normalization since the latter might be destabilized by small batch sizes. All training and evaluation processes were performed on a NVIDIA GeForce 2080Ti GPU and a 2.1GHz CPU.

### RNN-NDP+CA model

The proposed method combines RNN-NDP and CA model. We adopt abovementioned RNN-NDP model to calculate the suitability of lesion development, which is then used as an input parameter in CA model to simulate the development of lung infection in COVID-19 cases. **Figure S3** shows the flowchart of this model. This method consists the following steps. (1) Driving factors are extracted from the multi-stage CT scan data in the pre-treatment period. (2) A CNN is trained with the CT data and driving factor data sets. The suitability of the lesion development is calculated by the trained CNN. (3) The overall probability of lesion development is calculated by combining the suitability for lesion development, neighborhood effect and random factors. It is then used to simulate the data of various stages using CA model (similar to the RF+CA algorithm). (4) Considering the time interval and the lesion evolution speed based on the previous data, the final probability is estimated.

The network architecture is shown in **Figure S3** This model was trained and evaluated with the same configuration as that in the previous RNN-NDP model

### Alleviating the disadvantage of unequal interval among multi-stage CT examinations on the forecasting system

In the lesion forecast model, time interval plays a vital role, i.e. the progression of lesion region varies over different time intervals. Besides, the CT volumes from the same patient are collected at irregular time intervals. To introduce this information into account, the progression of lesion mask is embedded with the number of iterations, which is proportional to the actual time interval between two adjacent stages. Also, in the comparison method of RNN-NDP, intuitively, GRU (a variation of LSTM model) is used to describe the time interval information. Apart from this, the hypothesis of lesion development, i.e. the normal distribution over time, also considers the effect caused by time. In this way, the time interval information is used in proposed methods appropriately and the challenge that CT scans from the same patient is collected at different time interval is alleviated.

### Radiomics analysis - An application of the forecast system for lesion development system

#### Radiomics feature extraction

Radiomics features were extracted using *PyRadiomics* in Python[6]. A total of 1081 3D radiomics features were extracted to quantify lung and lesions information: 1) 266 first-order features reflecting the distribution of voxel intensities within the region of the lung; 2) 17 3D shape features describing the patient's lungs morphological and structural changes; 3) 798 texture features: gray level co-occurrence matrix (n=350); (4) gray level run length matrix (n=224); and (5) gray level size zone matrix (n=224). These features quantitatively distinguished the voxel-level difference between the lesion and non-lesion area. The optimal subset of 37 radiomics features associated with the severe illness of COVID-19 was selected by sequentially applying minimum-redundancy-maximum-relevance[7] and least absolute shrinkage and selection operator (LASSO) techniques[8]. Furthermore, we analyzed the dynamic development pattern of multi-phase CT images. The slopes of the dynamic time-varying curve of radiomics features on multi-phase CT images are calculated as follows:

$$\boldsymbol{S} = \frac{1}{N}\sum_{i=1}^{N} L_i S_i \tag{5}$$

$N$ represents the number of polylines segments in a curve, $L_i$ and $S_i$ represents the length and slope of the $i$th segment of polyline, respectively. The relative area is calculated using area under curve (AUC) of the dynamic time-varying curve of typical radiomics features on multi-phase CT images. First, the data is normalized according to the maximum value, e.g. for a sequence of 28-day radiological features, each value of the original features is divided by its maximum value in 28 days. The polyline is then redrawn, the relative area $\boldsymbol{A}$ is calculated as follows:

$$\boldsymbol{A} = \frac{A^{\circ}}{N} \times 100\% \tag{6}$$

$A^{\circ}$ is the normalized AUC of the redrawn polyline. $N$ represents the total days that the radiomics feature has been tracked.

#### Distinguishing mild and severe cases by dynamic analysis of major radiological characteristics

After voxel-level forecast for lesion development, we quantitatively analyzed the initial CT images and the dynamic development pattern of multi-phase CT images, respectively. The significance ranking of radiomic features were obtained for distinguishing between mild and severe illness of COVID-19. In the initial CT images, fourteen radiomic features are statistically significant (P<0.001), which contains one intensity feature (*original_firstorder_RootMeanSquared*), one shape feature (*original_shape_Sphericity*), twelve texture features (*original_glcm_Idmn*, *log-sigma-5-0-mm-3D_glcm_Contrast*, *wavelet-HHL_glszm_GrayLevelNonUniformity*, *wavelet-LLH_glcm_Imc2*, etc.), as shown in **Table S1**. Furthermore, in the dynamic development pattern of multi-phase CT images, we excavated the radiomic features with significant difference between mild and severe cases in terms of slope and area under the dynamic curve (AUC). The most significant difference in slope between mild and severe cases is found to be *Wavelet-HHL_gldm_DependenceVariance*, *original_shape_Sphericity* and other 11 features ($|S_{mild}-S_{severe}|>0.10$, $S_{mild}$ and $S_{severe}$ represent the slope of the time-varying curve for same radiological feature of the mild and the severe group respectively), as shown in **Table S2**. We ranked the top five features based on AUCs, and the ranks were consistent between the mild and the severe group. The slope's differences and AUCs of four radiological features had high values. However, the AUCs of one radiomic feature had an obvious difference between the mild and the severe group. Area of *original_glcm_Idmn* is 98.59% and 99.19%; Area of *log-sigma-2-0-mm-3D_glcm_Imc2* is 97.35% and 97.06%; Area of *log-sigma-1-0-mm-3D_gldm_DependenceEntropy* is 91.42% and 92.07%; Area of wavelet-*HHL_gldm_DependenceVariance* is 38.83% and 25.59%; Area of *original_shape_Sphericity* is 74.41% and 73.92%, as shown in **Figure S4a, b.**

**Table S1 | Radiomics features of the initial CT images from the patients with mild, severe illness (p < 0.001).**

| Radiomic feature | Severe (n=104) Mean (std) | Not Severe (n=180) Mean (std) | p-value |
|---|---|---|---|
| original_firstorder_RootMeanSquared | 721.40 (74.37) | 680.45 (67.80) | 0.000001 |
| original_glcm_Idmn | 0.98 (0.01) | 0.98 (0.02) | 0.000001 |
| log-sigma-5-0-mm-3D_glcm_Contrast | 1.89 (0.84) | 2.47 (1.32) | 0.000001 |
| wavelet-LLH_glcm_Imc2 | 0.83 (0.11) | 0.89(0.09) | 0.000001 |
| wavelet-HHL_glszm_GrayLevelNonUniformity | 98.78 (37.10) | 81.71(36.66) | 0.000001 |
| wavelet-HHH_gldm_DependenceEntropy | 4.76 (0.44) | 4.53 (0.49) | 0.000001 |
| original_shape_Sphericity | 0.18 (0.03) | 0.19 (0.03) | 0.000001 |
| log-sigma-2-0-mm-3D_glcm_Id | 0.51 (0.07) | 0.48 (0.06) | 0.000001 |
| wavelet-LLH_glszm_GrayLevelVariance | 46.64 (30.08) | 57.61 (31.83) | 0.000015 |
| wavelet-HLH_glcm_InverseVariance | 0.39 (0.09) | 0.42 (0.08) | 0.00007 |
| wavelet-HLH_glcm_JointEnergy | 0.08 (0.07) | 0.10 (0.08) | 0.000097 |
| wavelet-LHL_glszm_ZoneEntropy | 5.92 (0.16) | 5.87 (0.20) | 0.00029 |
| log-sigma-3-0-mm-3D_glszm_LargeAreaHighGrayLevelEmphasis | 48945.37 (82933.87) | 32014.05 (38340.12) | 0.000676 |
| log-sigma-1-0-mm-3D_gldm_DependenceVariance | 4.53 (3.94) | 3.638130 (2.61) | 0.000686 |
| wavelet-LLL_glcm_DifferenceAverage | 13.34 (4.69) | 14.48 (4.20) | 0.001402 |
| log-sigma-2-0-mm-3D_glcm_SumEntropy | 4.76 (0.38) | 4.65 (0.52) | 0.001895 |
| wavelet-LHH_glcm_Imc1 | -0.10 (0.06) | -0.12 (0.08) | 0.001988 |
| log-sigma-3-0-mm-3D_glcm_JointEnergy | 0.03 (0.01) | 0.03 (0.02) | 0.004486 |
| wavelet-LHH_glcm_DifferenceVariance | 8.10 (7.50) | 6.42 (7.48) | 0.005603 |
| wavelet-LHL_glcm_Correlation | 0.23 (0.28) | 0.29 (0.30) | 0.01013 |
| log-sigma-1-0-mm-3D_firstorder_Uniformity | 0.08 (0.03) | 0.08(0.02) | 0.014971 |
| log-sigma-2-0-mm-3D_glszm_SizeZoneNonUniformityNormalized | 0.26 (0.05) | 0.27 (0.05) | 0.016853 |
| wavelet-HLH_firstorder_Maximum | 334.35 (256.55) | 289.56 (270.69) | 0.036493 |
| wavelet-LHH_glszm_LargeAreaHighGrayLevelEmphasis | 27339.81 (76896.29) | 16785.37 (53647.54) | 0.043267 |
| log-sigma-1-0-mm-3D_gldm_DependenceEntropy | 6.48 (0.26) | 6.45 (0.25) | 0.061243 |
| original_glcm_InverseVariance | 0.20 (0.08) | 0.19 (0.06) | 0.148718 |
| wavelet-LLH_gldm_LargeDependenceLowGrayLevelEmphasis | 0.03 (0.08) | 0.038 (0.05) | 0.176577 |
| log-sigma-2-0-mm-3D_glcm_Imc2 | 0.96 (0.03) | 0.96 (0.03) | 0.262569 |
| wavelet-HLL_gldm_LargeDependenceHighGrayLevelEmphasis | 15707.56 (16152.38) | 14219.34 (17121.95) | 0.270441 |
| wavelet-HLH_gldm_LargeDependenceLowGrayLevelEmphasis | 0.84 (1.714979) | 0.97 (1.47) | 0.298007 |
| original_gldm_DependenceNonUniformityNormalized | 0.38 (0.14) | 0.37 (0.12) | 0.305104 |
| wavelet-HHL_gldm_DependenceVariance | 5.56 (6.14) | 5.18 (5.11) | 0.40134 |
| wavelet-LLH_gldm_DependenceNonUniformityNormalized | 0.27 (0.06) | 0.27 (0.05) | 0.418532 |
| wavelet-HLL_firstorder_Kurtosis | 14.46 (13.51) | 13.79 (12.04) | 0.508046 |
| original_shape_SurfaceVolumeRatio | 2.72 (0.64) | 2.74 (0.60) | 0.580828 |
| wavelet-LLH_firstorder_Range | 1156.46 (334.96) | 1169.24 (355.22) | 0.648126 |
| wavelet-HHL_glszm_LargeAreaHighGrayLevelEmphasis | 39779.33 (90517.10) | 38999.63 (102590.01) | 0.921217 |

#### Table S2 | The slopes of the dynamic time-varying curve of radiomics features on multi-phase CT images.

| Radiomics feature | Severe (n=104) | Not Severe (n=180) | Difference |
|---|---|---|---|
| wavelet-HHL_gldm_DependenceVariance | -0.02 | 2.04 | 2.06 |
| original_glcm_Idmn | 0.65 | 0.01 | 0.64 |
| wavelet-HLH_gldm_LargeDependenceLowGrayLevelEmphasis | 0.46 | -0.05 | 0.51 |
| log-sigma-3-0-mm-3D_glcm_JointEnergy | 0.73 | 0.28 | 0.45 |
| log-sigma-2-0-mm-3D_glcm_Imc2 | -0.42 | -0.01 | 0.41 |
| wavelet-LHH_glcm_Imc1 | -0.61 | -0.24 | 0.37 |
| wavelet-LLH_glcm_Imc2 | -0.19 | 0.01 | 0.20 |
| original_shape_Sphericity | 0.15 | 0.01 | 0.15 |
| log-sigma-1-0-mm-3D_firstorder_Uniformity | 0.28 | 0.16 | 0.12 |
| wavelet-LLH_gldm_LargeDependenceLowGrayLevelEmphasis | 0.40 | 0.29 | 0.11 |
| wavelet-HLH_glcm_JointEnergy | 0.16 | 0.05 | 0.10 |
| log-sigma-2-0-mm-3D_glcm_Id | 0.13 | 0.03 | 0.09 |
| original_gldm_DependenceNonUniformityNormalized | -0.17 | -0.09 | 0.08 |
| wavelet-LLH_gldm_DependenceNonUniformityNormalized | -0.19 | -0.11 | 0.07 |
| log-sigma-5-0-mm-3D_glcm_Contrast | -0.01 | -0.07 | 0.06 |
| wavelet-LLL_glcm_DifferenceAverage | -0.02 | 0.04 | 0.06 |
| wavelet-HLH_glcm_InverseVariance | 0.07 | 0.03 | 0.05 |
| log-sigma-1-0-mm-3D_gldm_DependenceVariance | 0.024 | -0.02 | 0.05 |
| wavelet-LHL_glszm_ZoneEntropy | 0.03 | 0.07 | 0.04 |
| log-sigma-2-0-mm-3D_glszm_SizeZoneNonUniformityNormalized | 0.02 | -0.01 | 0.03 |
| wavelet-LHH_glcm_DifferenceVariance | 0.003426 | -0.02 | 0.03 |
| log-sigma-1-0-mm-3D_gldm_DependenceEntropy | 0.000143 | -0.03 | 0.03 |
| original_glcm_InverseVariance | 0.12 | 0.10 | 0.02 |
| wavelet-LHL_glcm_Correlation | 0.03 | 0.02 | 0.02 |
| original_shape_SurfaceVolumeRatio | -0.03 | -0.04 | 0.01 |
| wavelet-LLH_glszm_GrayLevelVariance | -0.00486 | 0.01 | 0.01 |
| log-sigma-2-0-mm-3D_glcm_SumEntropy | -0.05 | -0.04 | 0.01 |
| wavelet-HLL_firstorder_Kurtosis | 0.02 | 0.02 | 0.01 |
| wavelet-HHH_gldm_DependenceEntropy | -0.02 | -0.01 | 0.002704 |
| wavelet-HHL_glszm_GrayLevelNonUniformity | -0.00053 | -0.00129 | 0.000763 |
| wavelet-HLH_firstorder_Maximum | -0.00021 | -0.00078 | 0.000571 |
| original_firstorder_RootMeanSquared | -7E-06 | 0.000031 | 0.000038 |
| wavelet-HLL_gldm_LargeDependenceHighGrayLevelEmphasis | -1.5E-05 | -5.3E-05 | 0.000038 |
| wavelet-LLH_firstorder_Range | -6.7E-05 | -4.3E-05 | 0.000024 |
| wavelet-HHL_glszm_LargeAreaHighGrayLevelEmphasis | -1.9E-05 | -6E-06 | 0.000013 |
| wavelet-LHH_glszm_LargeAreaHighGrayLevelEmphasis | -1.6E-05 | -9E-06 | 0.000007 |
| log-sigma-3-0-mm-3D_glszm_LargeAreaHighGrayLevelEmphasis | -5E-06 | -4E-06 | 0.000001 |

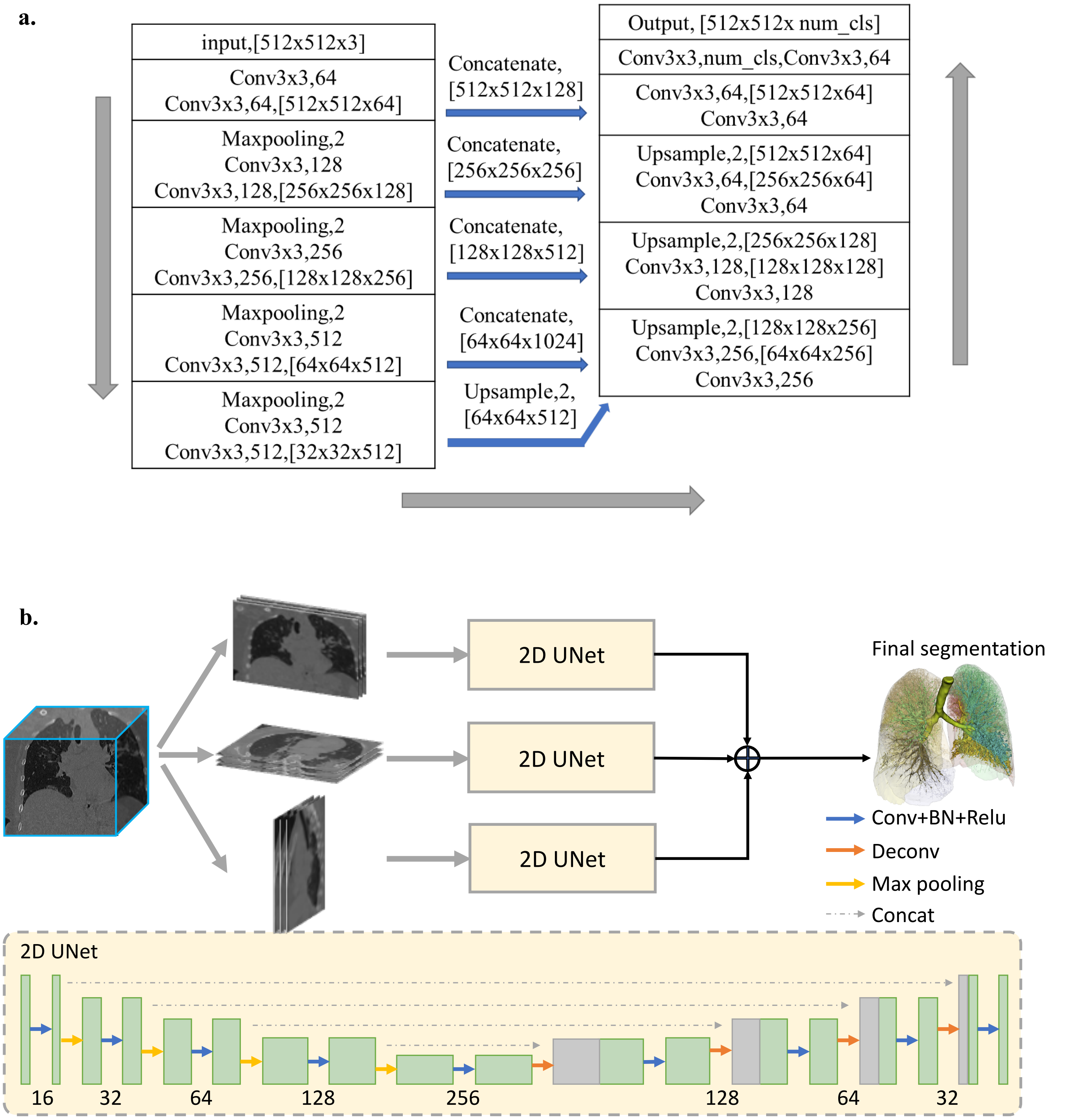


**Figure S1 | a.** 3D U-Net structure of lung segmentation network. **b.** Workflow of TAI segmentation in lung region using 2D U-Net. Both networks are based on U-Net, whose processing direction is U-like.

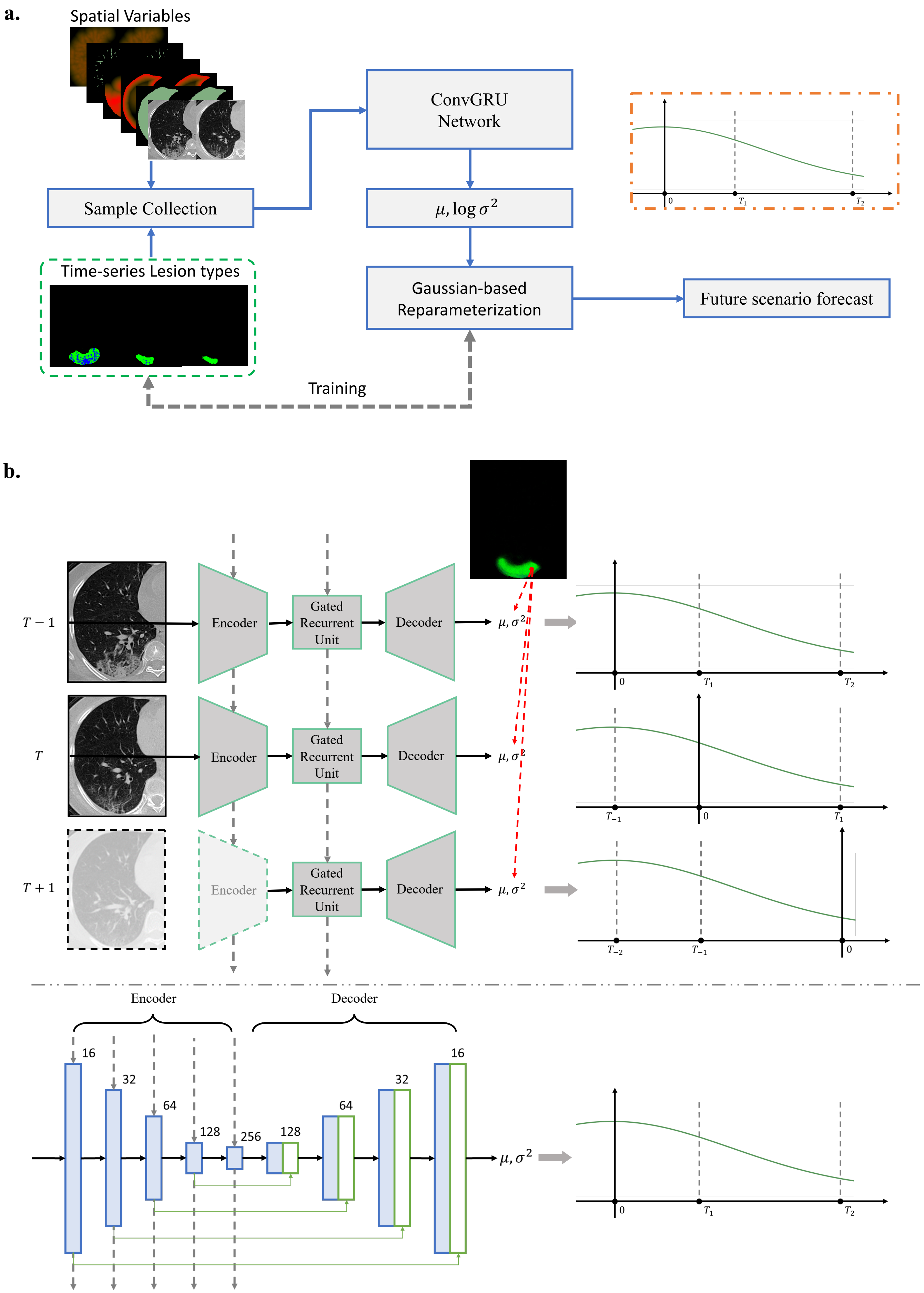


**Figure S2 | a.** Flowchart of lesion evolution by RNN-NDP model. **b.** Network architecture of Gaussian-based ConvGRU network used in RNN-NDP model.

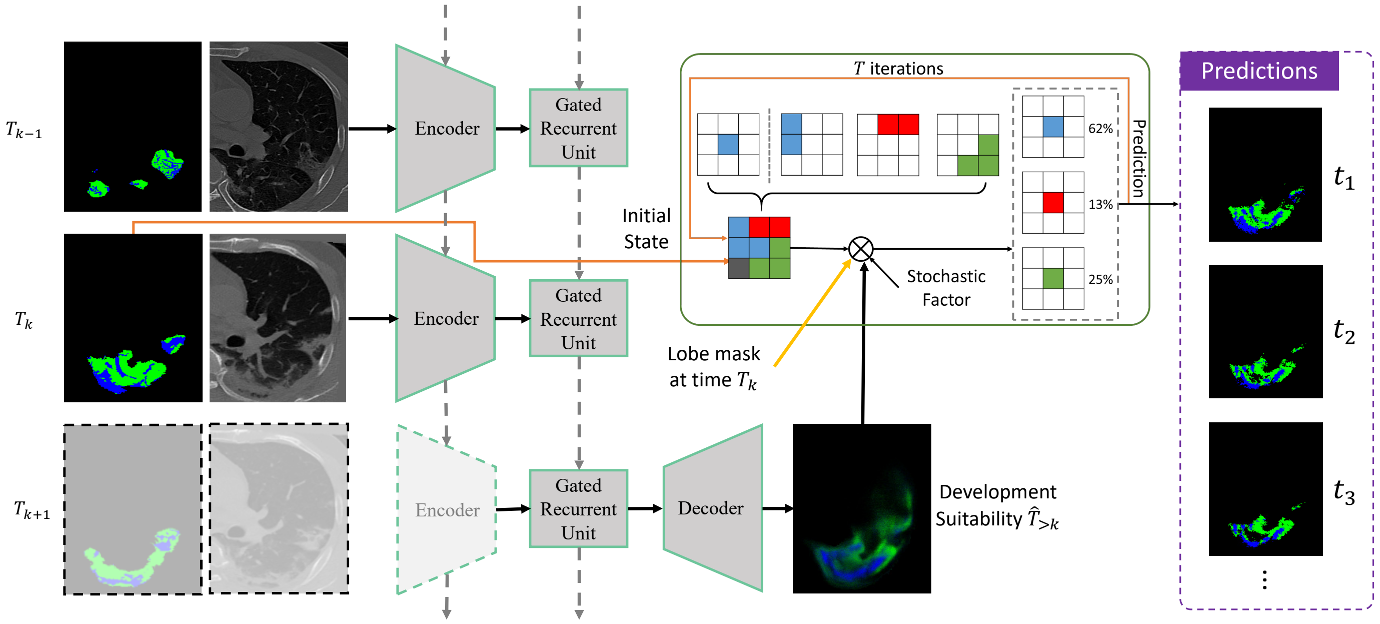


**Figure S3 | Forecast lesion evolution by RNN-NDP+CA model.** The output of network is regarded as development suitability.

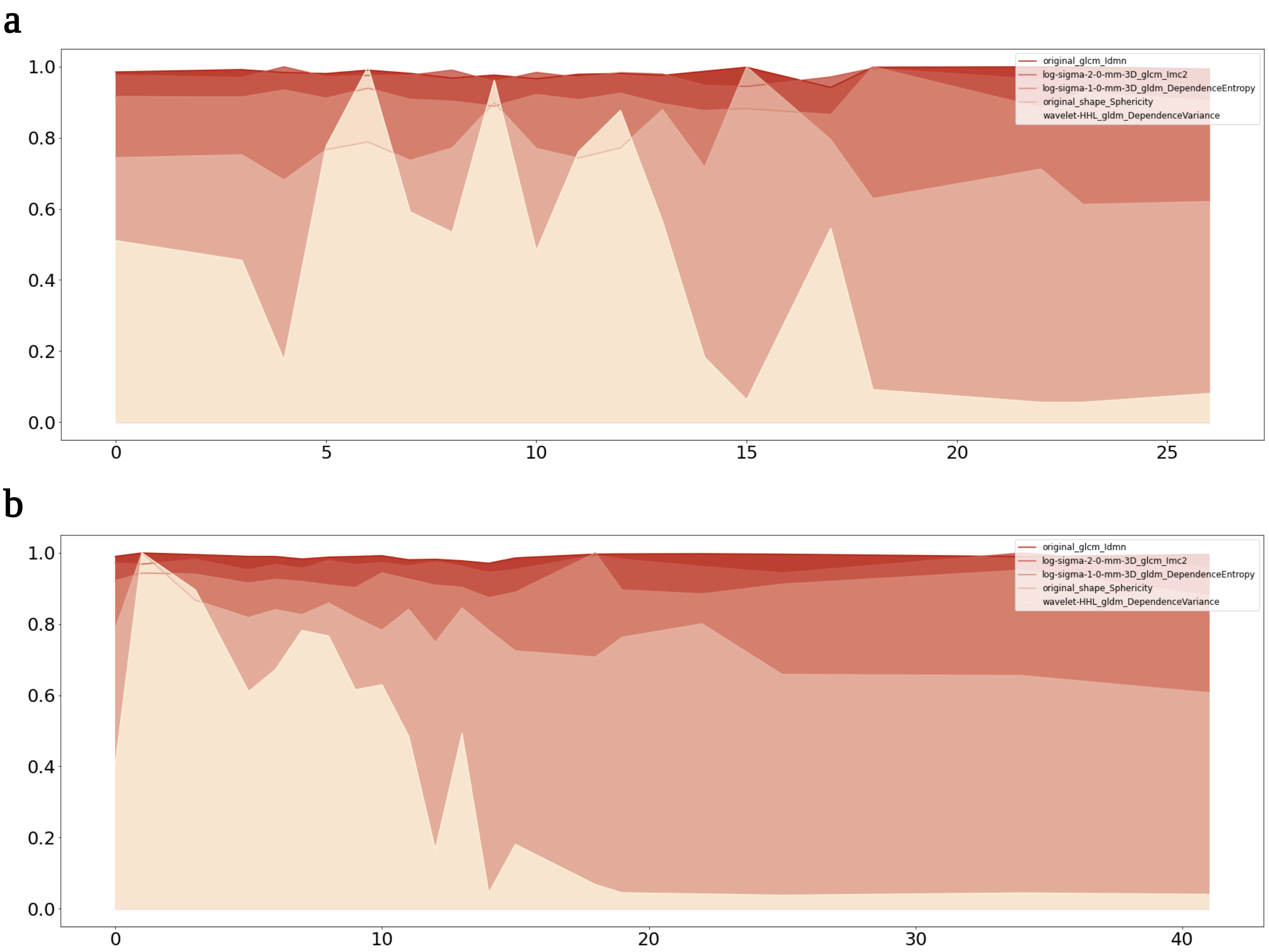


**Figure S4 | The AUC of the dynamic time-varying curve of typical radiomics features on multi-phase CT images. a.** mild illness of COVID-19. **b.** severe illness of COVID-19.
